# Combined prenatal *Lactobacillus reuteri* and ω-3 supplementation synergistically modulates DNA methylation in neonatal T helper cells

**DOI:** 10.1101/2021.03.25.21254287

**Authors:** Johanna Huoman, David Martínez-Enguita, Elin Olsson, Jan Ernerudh, Lennart Nilsson, Karel Duchén, Mika Gustafsson, Maria C Jenmalm

## Abstract

**Background:** Environmental exposures may alter DNA methylation patterns of T helper cells. As T helper cells are instrumental for allergy development, changes in methylation patterns may constitute a mechanism of action for allergy preventive interventions. While epigenetic effects of separate perinatal probiotic or ω-3 fatty acid supplementation have been studied previously, the combined treatment has not been assessed. We aimed to investigate epigenome-wide DNA methylation patterns in cord blood samples from children in a randomised double-blind placebo-controlled allergy prevention trial using pre- and postnatal combined *Lactobacillus reuteri* and ω-3 fatty acid treatment. To this end, >866 000 CpG sites (MethylationEPIC 850K array) in cord blood CD4+ T cells were examined in samples from all four study arms (double-treatment: n=18, single treatments: probiotics n=16, ω-3 n=15, and double placebo: n=14). Statistical and bioinformatic analyses identified treatment-associated differentially methylated CpGs and genes, which were used to identify treatment-induced network modules. Pathway analyses inferred biological relevance, and comparisons were made to an independent allergy data set.

**Results:** Comparing the active treatments to the double placebo group, most differentially methylated CpGs and genes were hypermethylated, suggesting induction of transcriptional inhibition. The double-treated group showed the largest number of differentially methylated CpGs, of which many were unique, suggesting synergy between interventions. Clusters within the double-treated network module consisted of immune-related pathways, including T cell receptor signalling, and antigen processing and presentation, with similar pathways revealed for the single-treatment modules. CpGs derived from differential methylation and network module analyses were enriched in an independent allergy data set, particularly in the double-treatment group, proposing treatment-induced DNA methylation changes as relevant for allergy development.

**Conclusion:** Prenatal *L. reuteri* and/or ω-3 fatty acid treatment results in hypermethylation and affects immune- and allergy-related pathways in neonatal T helper cells, with potentially

synergistic effects between the interventions and relevance for allergic disease. Further studies need to address these findings on a transcriptional level, and whether the results associate to allergy development in the children. Understanding the role of DNA methylation in regulating effects of perinatal probiotic and ω-3 interventions may provide essential knowledge in the development of efficacious allergy preventive strategies.

**Trial registration:** ClinicalTrials.gov, ClinicalTrials.gov-ID: NCT01542970. Registered 27^th^ of February 2012 – Retrospectively registered, https://clinicaltrials.gov/ct2/show/NCT01542970

## INTRODUCTION

Present-day increases in allergy prevalence in urbanised societies may be caused by the extensive loss of diverse microbial exposures due to changed lifestyle habits, thereby affecting development of appropriate immune responses.^1,2^ This is particularly important throughout the perinatal period, when maternal exposures during pregnancy and breastfeeding lay the foundations of, and influence, immune development of the foetus and the newborn child.^3-6^ Hence, the maternal influence on the foetal immune system is significant, and the composition of both the maternal diet and her commensal microbiome is crucial in the process.^7,8^

By supplementing pregnant women, with a high propensity of having an allergic child, during pregnancy and lactation with ω-3 fatty acids and probiotics, it may be possible to modulate the foetal immune system towards tolerance.^7,9,10^ In Westernised societies the dietary fatty acid intake favours ω-6 over ω-3 fatty acids ^11,12^, thereby promoting production of pro-inflammatory mediators, such as prostaglandins and leukotrienes ^13^, and possibly sustaining allergic inflammation.^10^ However, by ω-3 supplementation this ratio may be altered and promote downstream production of immunoregulatory mediators.^10^ ω-3 fatty acids may dampen allergic inflammation by inhibiting effects of IL-4 and IL-13, as well as by preventing isotype switching of B cells to IgE-antibodies.^7^ Indeed, prenatal ω-3 intervention attenuated IL-4 and IL-13 levels in cord blood^14^, and decreased the risk of developing asthma in the offspring.^15^ Previously, we have investigated the effects of pre- and postnatal ω-3 supplementation, demonstrating lower prevalence of sensitisation and IgE-associated eczema throughout the first year of life^16^, and a decreased risk of developing any IgE-associated disease up to two years of age^17^, in ω-3 compared to placebo-treated children. Several studies point at altered gut microbiomes in children developing allergies^18-23^, suggesting that increasing gut microbial diversity by *e*.*g*. probiotic intervention may be helpful in allergy prevention.^24^ Lactobacilli, a commonly used family of probiotic bacteria, induce both innate and adaptive immunostimulatory responses, apart from improving barrier integrity and modulating gut microbiota composition.^25^ We have previously shown allergy preventive effects on IgE-associated eczema upon supplementation with *Lactobacillus reuteri*.^26^ Interestingly, children whose mothers had allergic symptoms revealed the greatest benefits of the *L. reuteri* treatment^26^, whereas ω-3 treatment provided most benefit to children whose mothers did not have allergic symptoms.^16,17^ This suggests that combining the treatments could benefit allergy prone children regardless of maternal allergy status, and possible synergistic effects of the immunostimulating probiotics and the immunomodulatory ω-3 fatty acids could occur. However, this has thus far not been studied.

A suitable way to evaluate the effects of supplemental interventions would be to study epigenetic modifications, since they regulate transcriptional accessibility of genomic regions in response to environmental stimuli.^27^ DNA methylation of CpG sites is considered the most stable epigenetic modification, and is known to regulate the differentiation of T helper cells^28^, cells that are central in development of allergic disease.^27^ Indeed, differential DNA methylation patterns associate with development of sensitisation^29,30^, asthma^31,32^ and food allergies^33,34^ in children and adolescents. Epigenetic studies of prenatal fatty acid intervention have shown effects on histone modifications^35^, while others report conflicting results on DNA methylation patterning.^36,37^ A study on epigenomic modulation of combined *L. rhamnosus GG/Bifidobacterium lactis* in obesity^38^ revealed general hypomethylation, in line with findings from our previous study investigating *L. reuteri* in allergy prevention.^39^ In newborns whose mothers had been supplemented with *L. reuteri* throughout the last month of pregnancy, this loss of methylation in CD4+ cord blood cells was prominent in genes related to immune activation.^39^ This indicates that treated children received a head-start in their immune maturation, which hence may affect the development of allergic disease. However, in the context of combining probiotics and ω-3 fatty acids, neither clinical outcomes nor epigenetic effects have yet been studied.

In the on-going PROOM-3 trial (PRObiotics and OMega-3, ClinicalTrials.gov-ID: NCT01542970), the allergy preventive effects of combined *L. reuteri* and ω-3 fatty acids supplementation, from mid-pregnancy throughout the first year of life of the child, are studied. In this paper, we examined epigenome-wide DNA methylation patterns in cord blood CD4+ cells from the PROOM-3 cohort. Our hypothesis was that the earlier initiated probiotic treatment will lead to more prominent effects on DNA hypomethylation in immune-related genes compared to the previous study.^39^ Likewise, we hypothesised that ω-3 fatty acid supplementation will lead to hypomethylation of immune-related sites, and possibly work synergistically with the probiotic intervention to cause the greatest differential methylation in immunological pathways. Our findings indicate that the epigenome is modulated by the prenatal treatment with probiotics and ω-3 fatty acids, which may have implications for developing and improving future allergy prevention strategies.

## RESULTS

### Differential DNA methylation analyses reveal mainly hypermethylated CpGs and genes upon supplementation with *L. reuteri* and/or ω-3 fatty acids

Comparisons of demographic variables revealed no differences between the treatment groups, except for a higher proportion of females in the ω-3 single active treatment (Pω) group (Table 1). This lead us to exclude the sex chromosomes from further analyses, as retaining them might affect downstream results. Thereafter, we pursued multidimensional scaling (MDS) analyses to investigate inherent differences between the three active treatment groups, the double *L. reuteri* and ω-3-treated group (Lω), the *L. reuteri* single-treated group (LP) and the ω-3 single-treated group (Pω), compared to the double placebo group (PP). These analyses revealed distinct differences between the treated and untreated groups (Figure 1A-C), indicating treatment-associated changes at the epigenome level. We then proceeded to analyse differential DNA methylation, using an FDR-adjusted p-value threshold of <0.1 along with a mean methylation difference (MMD) of > 5% to define differentially methylated CpGs (DMCs). The double-treated Lω group demonstrated the highest number of DMCs compared with the single active treatment groups (Lω: 1659, LP: 1246 and Pω: 984 CpG sites, respectively, Table 2). A majority of the DMCs were hypermethylated (58-70%, Table 2, Figure 1 D-E, Table S1) in all three comparisons, suggesting that prenatal treatment with probiotics and/or ω-3 fatty acids foremostly leads to transcriptional inhibition. To investigate the biological impact of these findings, the DMCs were mapped to their corresponding differentially methylated genes (DMGs), which thereafter were utilised for pathway enrichment analyses. The double-treated Lω group contained 794 genes compared to 620 and 494 genes in the single-treated LP and Pω groups, respectively (Figure 2, Table S2), and 72 genes were shared for all three comparisons (Figure 2, Table S3). Pathway enrichment analyses of the DMGs in the respective comparisons revealed 87 pathways for the Lω group, including immune-related pathways such as PI3K-Akt and MAPK signalling,

**Table 1.**
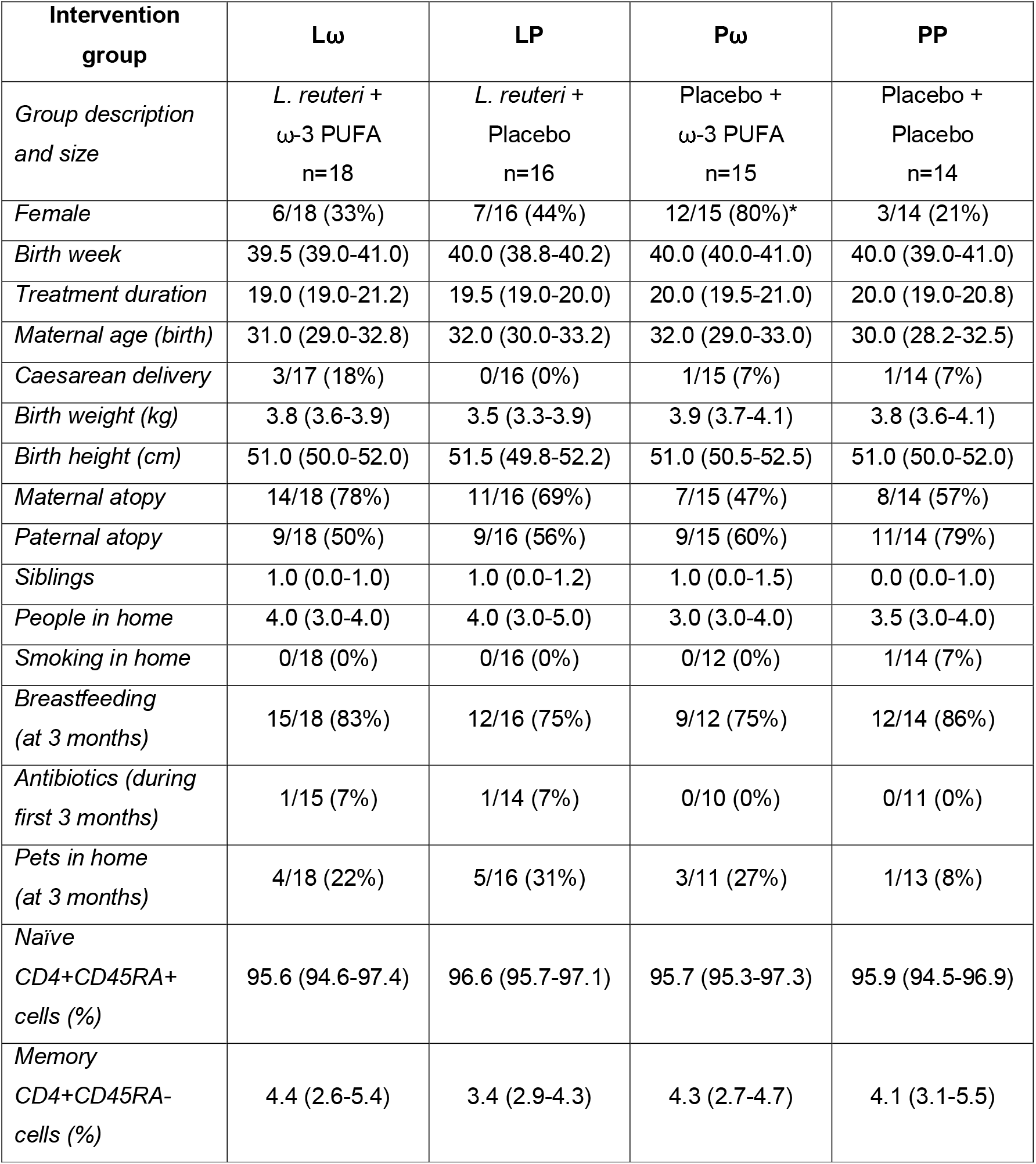
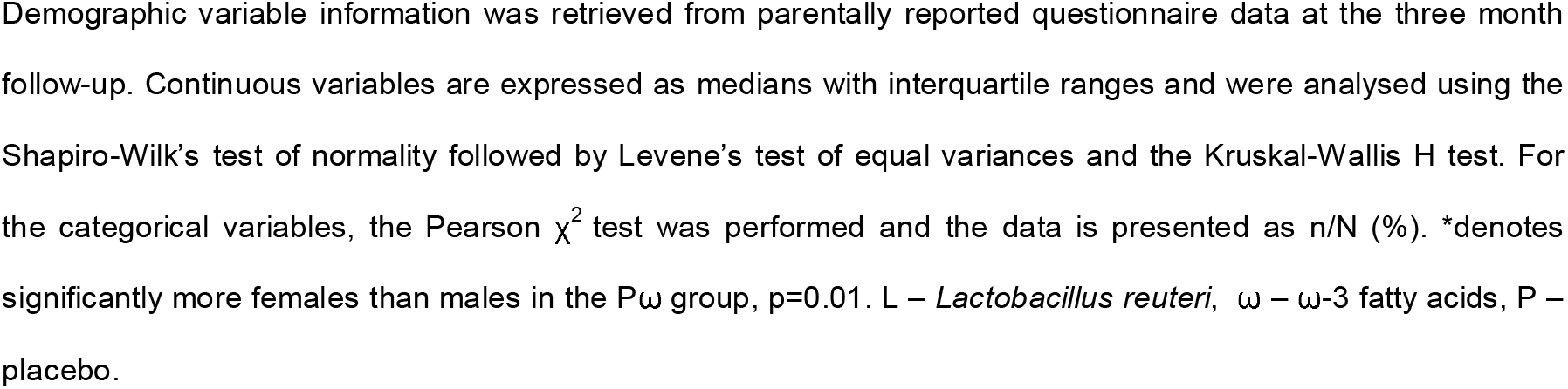
Demographic characteristics of children with available cord blood samples from the PROOM-3 study.

**Table 2.** Results from the differential methylation analyses, comparing the active treatments *vs*. the double placebo group.

| Intervention group | Lω vs. PP | LP vs. PP | Pω vs. PP |
| --- | --- | --- | --- |
| DMCs | 1 659 | 1246 | 984 |
| Hypomethylated CpGs | 576 (35%) | 380 (30%) | 415 (42%) |
| Hypermethylated CpGs | 1083 (65%) | 866 (70%) | 569 (58%) |
| DMGs | 794 | 620 | 494 |
| Hypomethylated genes | 247 (31%) | 204 (33%) | 184 (37%) |
| Hypermethylated genes | 524 (66%) | 405 (65%) | 295 (60%) |
| Mixed methylation genes | 23 (3%) | 11 (2%) | 15 (3%) |

Proportions of hypo- and hypermethylated CpGs and genes, from the differential DNA methylation analyses comparing the active treatment groups vs. the double placebo group, are presented as n (% of total). DMCs – differentially methylated CpGs, DMGs – differentially methylated genes, L – *Lactobacillus reuteri*, $\omega$ – ω-3 fatty acids, P – placebo.

**Figure 1.**
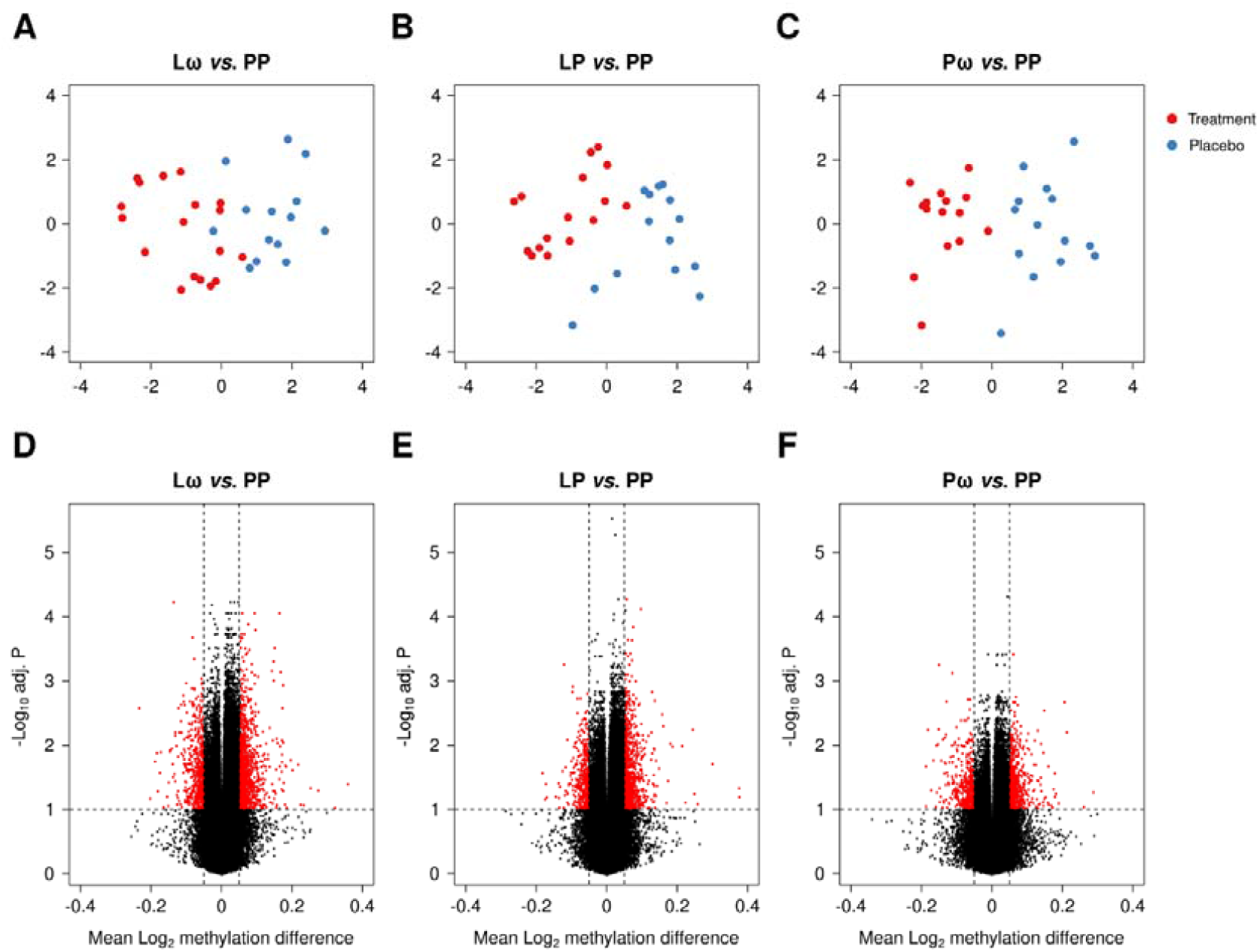
Graphical representations of multidimensional scaling (MDS) analyses are shown in panels A-C for the double active (Lω) and single active treatment groups (LP, Pω), respectively, in comparison to the double placebo group (PP). Red dots represent the active treatment group, blue dots the placebo group. In panels D-E, volcano plots illustrate the distribution of DMCs when comparing each of the active treatment groups to the double placebo group (PP). DMCs are defined as having an MMD of > 5% along with an FDR-corrected p-value of < 0.1, and are depicted as red dots. DMC – differentially methylated CpG, MDS – multidimensional scaling, MMD – mean methylation difference, L – *Lactobacillus reuteri*, ω – ω-3 fatty acids, P – placebo.

**Figure 2.**
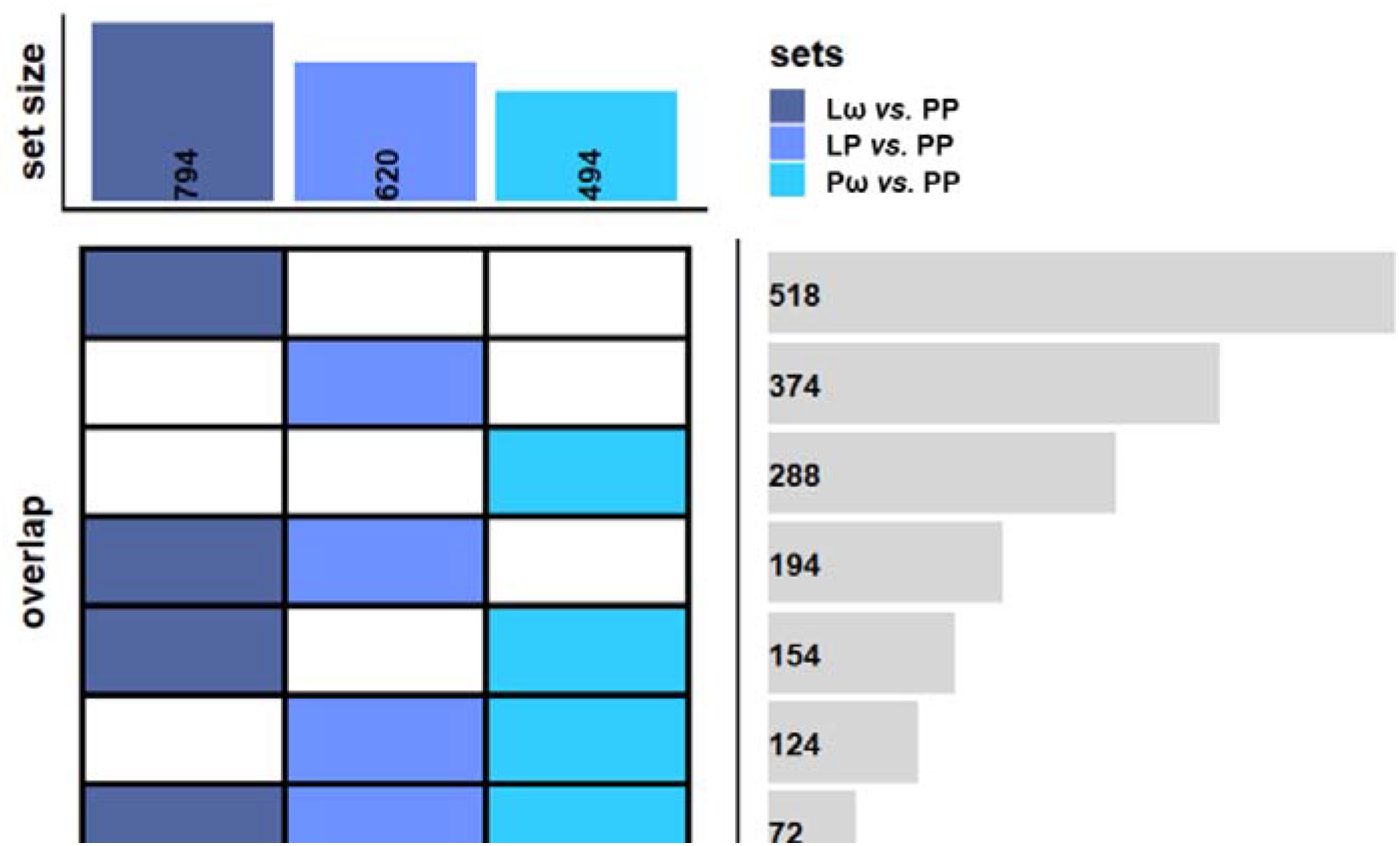
A graphic representation of overlapping DMGs between the different comparisons. The sets represent the different comparisons, and the intersectional sizes show the number of overlapping genes between the comparisons. Dark blue represents the double-treated group, light blue the *L. reuteri* single-treated group and coral blue the ω-3 single-treated group. L – *Lactobacillus reuteri*, ω – ω-3 fatty acids, P – placebo.

Phospholipase D signalling and T cell receptor signalling (Table S4). The LP group revealed 20 pathways, amongst others the Rap 1, Phospholipase D signalling and Th17 differentiation pathways (Table S5), whereas the Pω group did not reveal any pathways at all.

### Network analyses show immune-related pathways being affected by the interventions

As a means to further elaborate on the wider biological context in which the supplementation-induced DMGs act, the DMGs were mapped onto a protein-protein interaction map (STRING-db) to study putative interactions in a network context. The most interconnected modules were derived from DMGs identified by four module discovery methods from the MODifieR^40^ package. This approach narrowed down the number of supplementation-induced DMGs to 170, 100 and 87 genes for the Lω, LP and Pω comparisons, respectively (Tables S6-S8). High-confidence interactions from the respective modules are displayed in Figure 3 and Figures S2-3. Of the 170 genes identified in the double-treated Lω module, six genes were considered highly connected (exhibiting 10 or more connections to other module genes), and could be considered important hubs within the network: *FYN, GNB5, SNAP23, LAMC1, IL6* and *RAF1* (Figure 3). All genes but *IL6* were hypermethylated. Inspecting the network module for biologically relevant clusters, pathway enrichment analyses of those clusters revealed that supplementation with both *L. reuteri* and ω-3 affects pathways such as T cell receptor signalling, antigen processing and presentation as well as TGF-β signalling (Figure 3, Figure S3), clusters that were also observed in corresponding analyses of the single-treated groups (Figures S1-2, S4-5).

**Figure 3.**
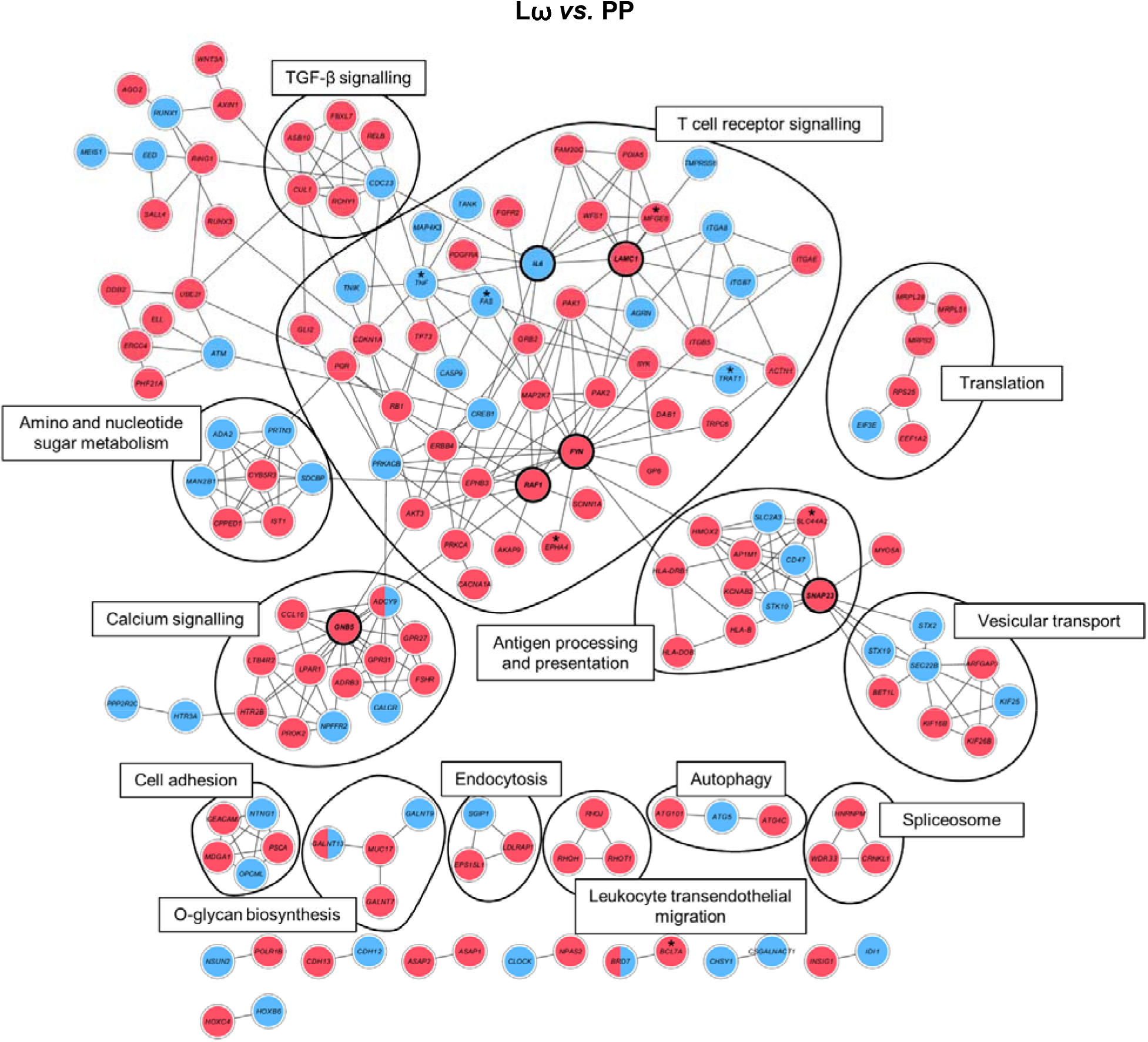
Visualisation of the consensus module created with the DMGs from the differential methylation comparison between the double-treated *L*.*reuteri*/ω-3 (Lω) and the double placebo (PP) group. Nodes represent genes and connecting lines represent protein-protein interactions (STRING combined score > 0.7) within the network. Red nodes illustrate hypermethylated genes, blue nodes indicate hypomethylated genes and both colours denote mixed methylation patterns. Black lines enclose biologically relevant clusters identified by bioinformatic pathway enrichment analyses. Nodes marked with * are also present in an independent allergy data set (described later in the paper). DMGs – differentially methylated genes, L – *Lactobacillus reuteri*, ω – ω-3 fatty acids, P – placebo.

To further evaluate common and unique features of the obtained supplementation modules, comparisons were made between the three treatment groups. While 10 genes were shared between all three comparisons, 24, 14 and 10 genes overlapped when comparing the Lω group to the *L. reuteri* single-treated (LP) group, the Lω to the ω-3 single-treated (Pω) group and the single-treated groups (LP *vs*. Pω) to each other, respectively (Figure 4A, Table S9). Genes common to all treatment groups related to cellular adhesion and extracellular matrix-receptor interaction pathways, whereas the comparisons between the single-treated groups revealed no significant pathways. Compared to the Lω double-treated group, overlaps to the single-active Pω group related to T cell receptor signalling and other related signal transduction pathways, while overlaps to the LP group included Ca2+ signalling and O-glycan biosynthesis

**Figure 4.**
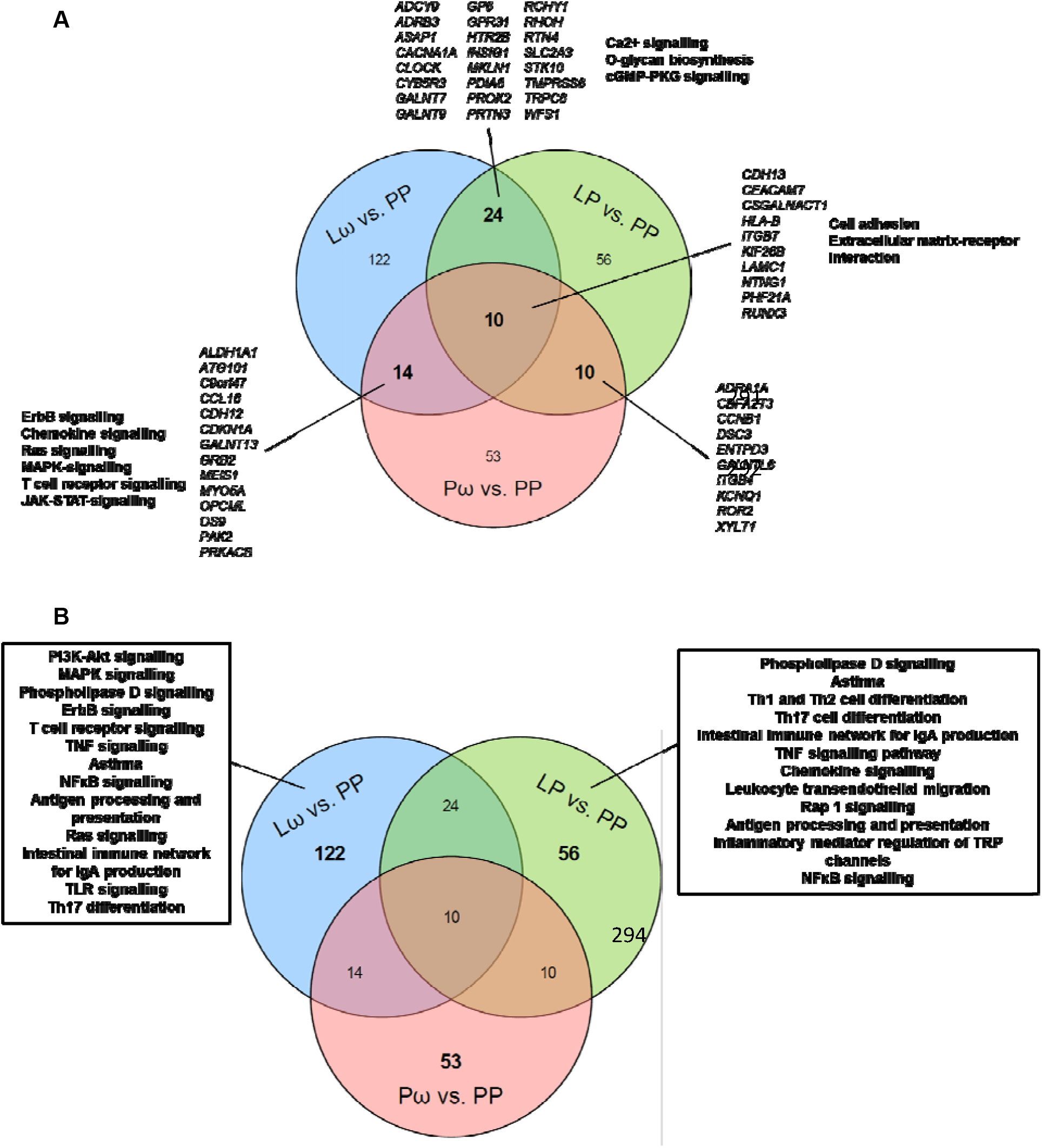
Venn diagrams illustrating the degree of overlap and uniqueness for the obtained genes from the three network modules for the double active treatment (Lω *vs*. PP) and the single active treatment (LP *vs*. PP, Pω *vs*. PP) comparisons. In panel A. the number of common genes from the overlap are displayed for each comparison is shown, with corresponding gene names denoted in italics, and selection of related pathways in bold. No relevant pathways were revealed for the shared genes of the LP *vs*. PP and Pω *vs*. PP comparisons. In panel B. the number of genes that are unique for each comparison are displayed along with a selection of corresponding related pathways. No pathways of relevance were revealed for the unique genes from the Pω *vs*. PP comparison. L – *Lactobacillus reuteri*, ω – ω-3 fatty acids, P – placebo.

(Figure 4A), pathways which are relevant for proper T cell function. The largest number of unique genes were revealed for the double active Lω treatment group (122 genes), which were mainly biologically related to T cell receptor signalling and related signal transduction pathways, TNF and NFκB signalling as well as asthma (Figure 4B). The single-treated groups showed similar numbers of unique genes (LP: 56 genes, Pω: 53 genes), which related to Th1/2/17 helper cell differentiation and asthma amongst other pathways for the LP group, while yet again no relevant pathways were discovered for the Pω treatment group. Hence, combined probiotic and ω-3 supplementation seems to have synergistic effects at the network level compared to the single treatments.

Further evaluation of the direction of methylation in the shared genes revealed that treatment with either *L. reuteri* or ω-3 fatty acids led to consistent methylation patterns in most of the genes when compared to the double-treated Lω group (Figure 5A-B, Table S9). This pattern was even more distinct comparing the single active treatment groups, where only one gene revealed different methylation patterns between the groups (Figure 5C). In the comparison between all three treatment groups, however, the patterning of approximately half the genes were similar, whereas the single active groups exhibited different and still distinct shared DNA methylation patterns with the Lω group (Figure 5D).

**Figure 5.**
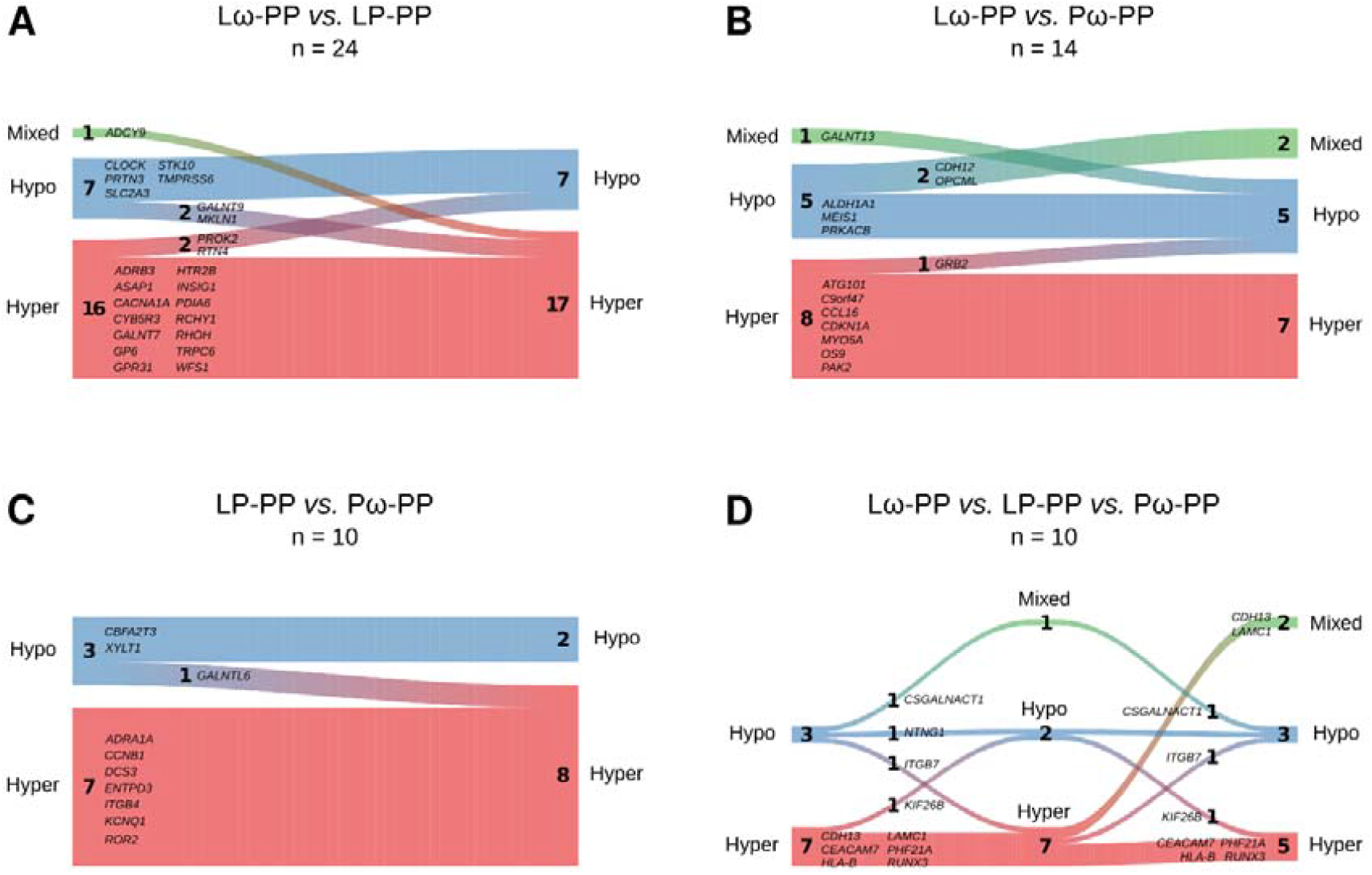
River plots illustrating the methylation status for overlapping genes being shared between the double-treated Lω group and A. the single-treated probiotic (LP *vs*. PP), and B. ω-3 treated (Pω *vs*. PP) group. Panel C. illustrates comparisons between the single treated groups (LP and Pω *vs*. PP) and D. shows the comparison between all treatment groups. Genes are represented as flows of size proportional to number of genes within their methylation status in a particular set, and differences between sets are shown as connections between flows. The respective gene names are denoted next to each flow. Red represents hypermethylation, blue hypomethylation and green a mixed methylation pattern. The number of genes in the respective flows are denoted in the graph. L – *Lactobacillus reuteri*, ω – ω-3 fatty acids, P – placebo.

### Differentially methylated CpGs in the active treatment groups were enriched in an independent allergic disease data set

As no information is as yet available on allergy development for the children included in this study, we pursued enrichment analyses on DMCs extracted from the differential DNA methylation analyses for all three comparisons, and compared them to DMCs of a publicly available allergy data set, studying DNA methylation patterns in CD4+ T cells of adults with seasonal allergic rhinitis (8 patients *vs*. 8 controls). Indeed, for all three comparisons, a substantial number of DMCs were significantly enriched within the allergy data set (Lω: 91, p=4.7e^-28^, LP: 47, p=2.6e^-11^ and Pω: 56 CpGs, p=1.9e^-17^, Figure 6A). Investigating involved pathways of the genes from these DM.Ce-s, few or no pathways emerged (results not shown). Three CpG sites overlapped between the three groups (in the genes *MYOF, C6orf123* and *PTPRN2*), while additionally 8 and 11 CpGs were common comparing the the double-treated Lω group to the single-treated LP and Pω groups, respectively, and 4 CpGs were overlapping between the single-treated groups (Table S10).

**Figure 6.**
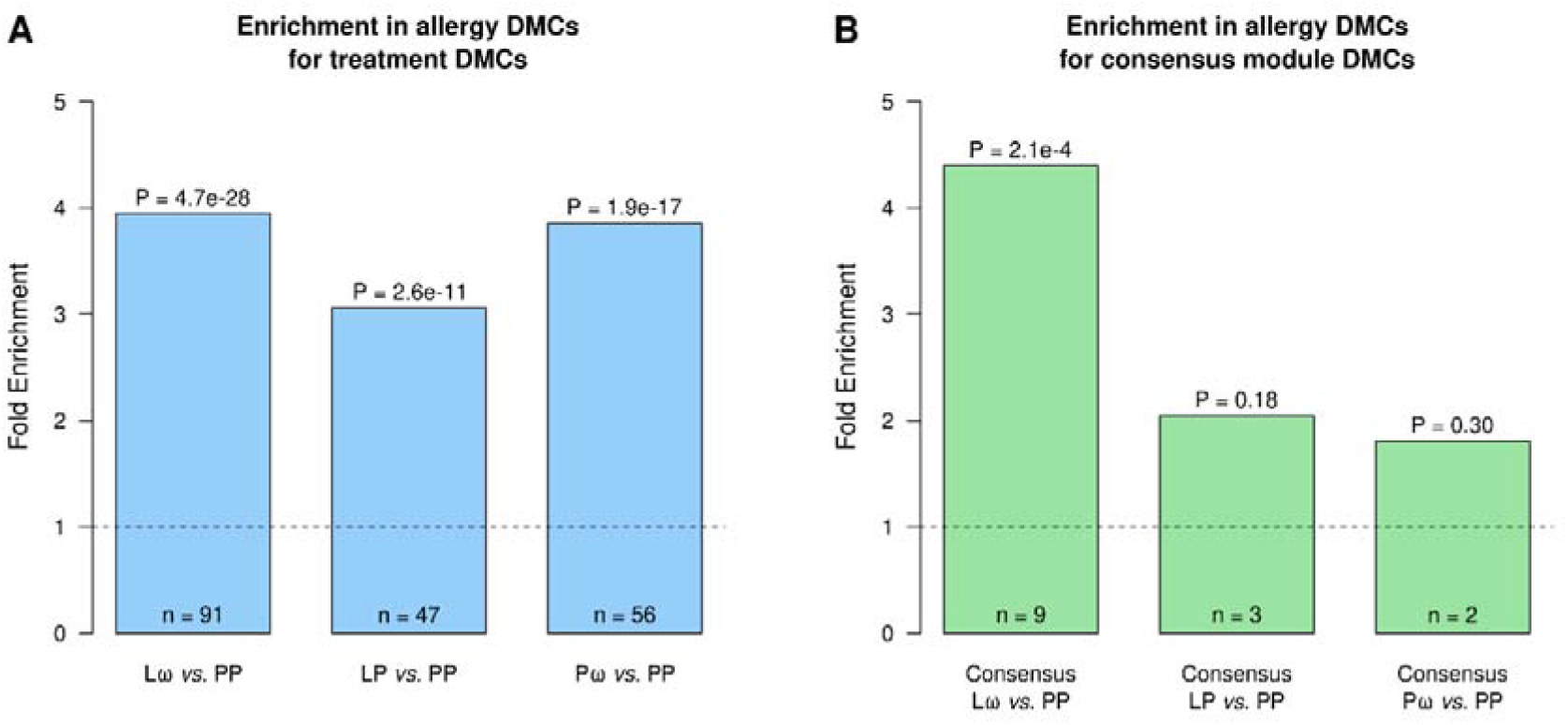
Enrichment analyses relating DMCs from A. differential DNA methylation analyses and B. network analyses for all three comparisons, to DMCs from a study investigating DNA methylation patterns in adults with seasonal allergic rhinitis. Panel A. shows treatment DMC overlaps with allergy DMCs. Panel B. shows consensus module gene associated DMC overlaps with allergy DMCs. The number of overlapping DMCs is shown at the bottom of the bars (n). A one-sided Fisher’s exact test was used for the enrichment analyses. Computed P-values represent the enrichment significance, while the fold enrichment illustrates how over-represented the overlap is, compared to what would be expected by chance for the given background.

As some results from the network analyses indicated involvement of network genes in allergy-related pathways, we additionally pursued comparisons of our DMCs from the consensus modules for all three comparisons to the above mentioned allergy data set. Enrichment analyses revealed that there was a significant overlap between our DMCs identified by the double-treated Lω network module and allergic disease (9 genes, p=0.00021, Figure 6B), while no significant enrichments were revealed for the single-treated groups. Examining these nine genes within the Lω module in more detail, four were hypomethylated (*TNF, FAS, TRAT1* and *HIVEP3*), while five were hypermethylated (*SLC44A2, MFGE8, EPHA4, BCL7A* and *RAB11FIP1*), and seven of these genes were present within the connected components of the network (all but *HIVEP3* and *RAB11FIP1*, Figure 3, genes marked with asterisks). Six out of seven genes were furthermore located to the central parts of the network, mainly involved in T cell receptor signalling and antigen processing and presentation. Comparing the direction of methylation to the independent allergy data set, four genes exhibited the opposite direction of patterning (*HIVEP3,TRAT1, BCL7A* and *SLC442A*), while the DNA methylation patterning of the remaining genes were identical between the groups (Figure S6).

## DISCUSSION

The role of perinatal combined allergy prevention interventions on epigenetic patterning in T helper cells has hitherto been unknown. In this paper, we set out to assess the epigenome-wide effects on cord blood CD4+ cells of combined maternal probiotic and ω-3 intervention during the latter half of pregnancy. Indeed, we show that prenatal supplementation affects the epigenome of T cells from cord blood in the neonates substantially, and differently so depending on the treatment combination.

The main finding of this study was that the largest treatment-induced effect on the epigenome was revealed in the Lω double active treatment group, and that regardless of treatment hypermethylation was the main outcome. The network analyses revealed the same pattern in the created intervention modules, and the genes shared between the comparisons as well as the performed network analyses revealed common biological pathways. The Lω module made up the highest number of genes while fewer, but still significant, numbers were revealed in the single-treated LP and Pω modules. This indicates that there might be some synergistic effects of the double treatment. Although the number of shared genes from the networks were limited and the methylation patterns of the genes were only partly similar, this further suggests that the combined treatment may modulate the epigenetic response differently from the single treatment groups. Probiotics may affect DNA methylation either by the production of metabolites downstream of short chain fatty acids, which are indispensable co-factors for proper functioning of enzymes such as DNA methyltransferases, or directly by modifying the actions of the same enzymes.^41^ Similarly, ω-3 fatty acid intervention may alter DNA methyltransferase activity^42^ and regulation of transcription factor activity.^43^ However, how the combination of the two treatments act on a molecular level still needs further elaboration. Examining the genes that were unique to the respective comparisons, the Lω and LP groups featured large numbers of immune-related pathways, yet again involving signal transduction pathways important for T cell signalling, but also T helper cell differentiation and asthma. This is particularly interesting, and future studies on these children will have to settle whether induced DNA methylation differences are associated with the development of allergic disease. Altogether, our findings propose that the interventions modulate the epigenome, and more specifically so in pathways that are of general importance for proper T cell development and function.

To further understand the impact of the prenatal intervention on the epigenome, genes that were highly connected within the module network were scrutinised in detail. In the double-treated Lω group, six genes had more than 10 connections to other genes within the network. One of the genes was *FYN*, a non-receptor tyrosine-protein kinase involved in the downstream signal transduction pathway of the T cell receptor, regulating amongst other things T cell activation and differentiation.^44^ In this study, *FYN* was hypermethylated, suggesting that the gene would be transcriptionally silenced by treatment and thereby also downregulating putative protein functions. Another gene of interest was the hypomethylated inflammatory cytokine gene *IL6*. IL-6 is heavily involved in the development and maintenance of inflammation, by inducing both innate and adaptive inflammatory immune responses.^45^ In modulation of the latter, IL-6 regulates both proliferation and cytokine responses of CD4+ cells by inducing JAK/STAT, MAPK and PI3K-Akt signalling pathways, and downstream effects of this activation have been implicated in development of asthma. On an epigenomic level, hypomethylation of IL-6 has been shown in children with peanut allergy^46^, and it has also been associated to worse lung function in asthmatic children.^47^ Furthermore, in the first 6-12 months of life, the *IL6R* gene was hypomethylated in a pre-and postnatal probiotic intervention study investigating development of obesity in children.^38^ As we are still collecting information about development of allergic disease such as asthma in the present PROOM-3 study, the long term implications of this finding are unknown. It is possible, however, that the intervention enables transcriptional accessibility of this gene, and hence equips the child with early inflammatory capacity. This notions is in line with the observed overlap of DMCs from the Lω consensus module with the publicly available allergy data set. Three hypomethylated genes from the T cell receptor signalling cluster were of particular interest: *TNF, FAS* and *TRAT1*. The T cell receptor-associated transmembrane adaptor protein TRAT1 is known to stabilise the T cell receptor/CD3 complex, and may thereby modulate downstream signal transduction and promote T cell function.^48,49^ TRAT1 was hypomethylated in the Lω group, whereas it was hypermethylated in the independent allergy data set, indicating that the combined intervention may enable proper T cell signalling propagation into the cell. While the cytokine tumor necrosis factor (TNF) may not only promote inflammatory responses and induce proliferation of both naïve and memory T cells, it may also drive apoptosis of effector T cells.^50^ Similarly, the TNF-related FAS receptor may also induce apoptosis of cells by binding to FASL, leading to recruitment of caspase-8 and −10 and subsequent commencement of programmed cell death.^51^ This process is of crucial importance in the establishment of peripheral tolerance, and as TNF and FAS were hypomethylated in the Lω comparison, this suggests that anti-inflammatory effects of TNF and FAS could be promoted by the supplementation of probiotics and ω-3 prenatally. However, further studies on transcriptional and protein levels of these mediators, along with functional studies in relation to allergy development should shed light on the relevance of these findings upon prenatal interventions.

One quite surprising finding of this study was that, in contrast to our hypothesis and results from our previous epigenome-wide study on the effects of perinatal *L. reuteri* treatment on cord blood CD4+ cells^39^, a larger proportion of the CpG sites were hypermethylated when comparing either of the intervention groups to the double placebo group. This finding could have several explanations. First of all, unlike our previous studies investigating possible allergy preventive effects of perinatal probiotic intervention^39^ or ω-3 intervention^16,17^ the combined intervention was initiated already at gestational (GW) 20 in the PROOM-3 study, compared to GW 25 for ω-3 fatty acid^16^ and GW 36 for *L. reuteri*^52^ supplementation in the previous study. Additionally, the maternal dosage of the *L. reuteri* strain was also higher by an order of magnitude in this study (10^9^ colony forming units (CFU), compared to 10^8^ CFU previously^52^), possibly allowing for potentiated effects of the probiotic intervention. Furthermore, we here utilised Illumina’s Infinium MethylationEPIC 850K array, surveying more than 866 000 sites all over the genome. Despite including more than 90% of the investigated sites from the previous Illumina 450K platform^53^, almost twice the amount of DNA methylation probes were investigated. This could possibly explain why the present findings differ from the previous study where the Illumina 450K platform was used. Also, as only autosomal chromosomes were retained the present analyses, this may have affected the outcomes in relation to our previous study^39^, where all chromosomes were included in the analyses. Altogether, these modifications to the study, experimental and statistical protocol could possibly explain the apparent reversed observations in terms of major hypermethylation upon intervention with *L. reuteri* and/or ω-3 fatty acids in this study, in contrast to hypomethylation upon *L. reuteri* intervention previously.^39^

Advantages of the present study include the performance of the DNA methylation analyses in purified CD4+ T cells, cells which are instrumental for development of specific long-term immune responses in general, but also for the development of allergic diseases in particular.^2^ Also, despite our relatively small sample numbers, we are able to evaluate all four intervention groups in this study, enabling us to shed light on treatment-specific effects on the methylome. The performance of network analyses provides another benefit of this study, where the analyses have generated putative candidate genes to study further in an allergy development and prevention context. As the study is still on-going, we were not able to perform any comparisons of the interventions and their respective DNA methylation patterns on outcomes of allergy development. However, we made up for this drawback by comparing our findings to an independent allergy study, where we indeed could show enrichment of our DMCs. Another limitation in our analyses include the lack of validation at the mRNA or protein expression level.

In conclusion, we demonstrate that the combined prenatal treatment with *L. reuteri* and/or ω-3 fatty acids results in general differential hypermethylation in neonatal CD4+ T cells, which affects both T cell-specific and immune-related pathways. This was corroborated by network analyses that condensed the findings to differentially methylated genes in linked pathways, which also showed relevance in an independent cohort of allergic disease. Future follow-ups on allergic disease and studies on transcriptional activity will confirm the long-term relevance of the presented findings, which may have an impact on future development of allergy prevention regimens.

## METHODS

### Study participants

In this study, cord blood samples collected at the time of birth were included for 63 children from the on-going randomised double blind placebo-controlled trial PROOM-3 (ClinicalTrials.gov-ID: NCT01542970), in which the allergy preventive effects of combined pre- and postnatal *Lactobacillus reuteri* and ω-3 fatty acid supplementation are investigated. The study is four-armed comprising the following groups; double active treatment (Lω, *L. reuteri* and ω-3 fatty acids), single active treatments (placebo for either treatment, active treatment for either *L. reuteri* (LP) or ω-3 (Pω)) and double placebo treatment (PP). In the present study, the following numbers of samples were available per group: Lω = 18, LP = 16, Pω = 15, PP = 14. The mothers are supplemented with the probiotic intervention or the corresponding placebo supplement from GW 20 until delivery, whereas the children receive the same treatment throughout the first year of life. The active probiotic substance consists of *L. reuteri* (BioGaia^®^, Stockholm, Sweden) suspended in sun flower and palm kernel oil, which is given as droplets twice daily to the mothers (10^9^ CFU) and once daily to the children (10^8^ CFU). Placebo treatment for the probiotic intervention consists of the same oil mixture as above without probiotics. The ω-3 fatty acid intervention or corresponding placebo supplement are given from GW 20 throughout lactation to the mothers. Twice daily, the mothers ingest either three capsules of Pikasol^®^ (Orkla Health, Lund, Sweden, 1 g capsules containing 640 mg ω-3 PUFA, 35% EPA, 25% DHA) or the placebo supplement olive oil. The children are followed-up at several occasions using questionnaires and by performance of clinical examinations. As the study still is ongoing, only parent reported questionnaire data from the 3-month follow-up were included for the purpose of this study. Parents or legal guardians provided informed consent on behalf of the children prior to inclusion in the study. Ethical permission for this study has been granted by the Regional Ethics Committee for Human Research in Linköping (Dnr 2011/45-31).

### Cord blood mononuclear cell and CD4+ magnetic bead isolation

The isolation of cord blood mononuclear cells (CBMCs) from cord blood and the following magnetic bead isolation of CD4+ T cells from the CBMCs have been described in detail elsewhere.^39^

### Flow cytometry measurements of naïve and memory T cell proportions

In order to determine the proportion of naïve and memory cells from the isolated CD4+ cells 0.2 × 10^6^ cells were stained for flow cytometry. Firstly, in order to determine the amount of live and dead cells in each sample Aqua LIVE/DEAD^™^ Fixable Dead Cell Stain (Invitrogen, Eugene, OR, USA) reconstituted reagent (according to manufacturer’s protocol) was diluted 1:500 in PBS (Medicago AB, Uppsala, Sweden) + 0.1% FCS (Sigma-Aldrich, Ayrshire, United Kingdom) and added to the samples for 15 minutes at RT. Thereafter, the cells were further stained with CD4 PerCP (SK3) and CD45RA FITC (L48), with a complementary isotype tube including CD4 PerCP and Mouse γ1 FITC (X40) (all antibodies: Becton-Dickinson, San José, CA, USA) for 15 min at 4°C. After washing the cells, they were resuspended as described previously^39^ and 10 000 events were collected on a FACS Canto II instrument using the FACS Diva software (version 8.0.1). The flow cytometry data were analysed using Kaluza (version 2.1, Beckman Coulter, Indianapolis, IN, USA), and the utilised gating strategy is presented in Figure S7.

### DNA extraction

From the remaining CD4+ cells, DNA was extracted using Allprep RNA/DNA Mini Kit (Qiagen, Austin, TX, USA) according to the manufacturer’s instructions. DNA concentrations were measured using Qubit dsDNA BR Assay Kit (Thermo Fischer Scientific, Waltham, MA, USA) according to the manufacturer’s instructions on a Qubit 3.0 fluorometer device (Thermo Fischer Scientific, Waltham, MA, USA). DNA samples were sent to the SNP&SEQ-technology platform at SciLifeLab (Uppsala University, Uppsala, Sweden) for the performance of Illumina’s Infinium Human MethylationEPIC 850K bead chip array preceded by bisulphite conversion on site. The amount of DNA for each sample ranged from 1000-1800 ng.

### Statistics

As the study is still on-going, the study has not yet been unblinded. An independent statistician has provided the code key only to collaborators that have not been involved in sample collection or laboratory experiments, as not to bias the investigators in interpretation of the data.

Descriptive analyses of demographic variables were performed. Continuous variables were tested for normality using the Shapiro-Wilk test of normality followed by Levene’s test of equal variances. Thereafter the Kruskal-Wallis H test was performed to study differences in proportions between the groups. For the categorical variables, the Pearson χ^2^ test was performed.

### DNA methylation analyses

The generated raw data from the Illumina 850K MethylationEPIC arrays were processed in GenomeStudio (version 2011.1, Illumina Inc.) by the SNP&SEQ-technology platform, to render IDAT-files manageable for downstream analyses in the R programming environment (version 3.6.3).

### Pre-processing and quality control

The raw DNA methylation data were pre-processed using the ChAMP^54^ package (version 2.16.1) for the R programming environment. Probes with detection p-values above 0.01 and multi-hit probes were filtered out, as were non-CpG probes and probes related to SNPs and NA-probes. Furthermore, as we found significantly more females in the single active ω-3 treated (Pω) group, and as X-inactivation in females skews the beta value distribution (Figure S8), probes located on the X and Y chromosomes were omitted from the data. After the filtration step, 738 451 CpGs of the initial 865 918 investigated CpGs in the Illumina MethylationEPIC 850K array remained. Thereafter, beta values were calculated for all probes, representing the mean methylation of each CpG site, and to adjust for the inherently different designs of the Illumina DNA methylation arrays, normalisation of the beta values was conducted using the beta mixture quantile (BMIQ) method.^55^ In order to identify the most significant components of variation in the data, and correct for unwanted variation if found, singular value decomposition (SVD) analyses were performed on the filtered and normalised data, using the ChAMP package. Based on results from these analyses, corrections were made in the data for the technical factors array and slide (Figure S9). The quality of the data was examined prior to and after the normalisation step (Figure S10). To ascertain possible differences in cellular content of the included samples, several methods were used to validate the findings. Maternal contamination of the cord blood samples was examined on the unfiltered data using the method proposed by Morin *et al*.^56^ By surveying threshold DNA methylation levels of 10 specified CpG sites, samples with values above the threshold values at five or more sites indicate contamination and should be excluded. Six samples had a beta value above the threshold value at one CpG site and one sample had beta values above the threshold values at two CpG sites (data not shown), indicating no maternal contamination and hence all 63 samples were retained in further downstream analyses. The cell type composition of the samples was evaluated on filtered and normalised samples using the Houseman method.^57^ The majority of cells were CD4+ cells with median proportions in the intervention groups between 94-96% (Table S11), indicating high sample purity and successful isolation of CD4+ T cells. Multidimensional scaling (MDS) analyses were performed on the filtered, normalised and batch corrected data to study inherent differences in the data, using the R package minfi (version 1.32.0). Results of the SVD analyses using the ChAMP package showed that, apart from the sample array and slide, sex was a major source of unwanted variation in the normalised data. Therefore, sex was added as a co-variate in the batch correction step for the data used in the MDS analyses, using the ComBat function^58^ from the sva package^59^ (version 3.34.0), implemented in ChAMP. Following the statistical analyses, the MDS results were graphically represented by assigning the samples to their respective intervention group belonging.

### Differential DNA methylation analysis

Using the limma package (version 3.42.2), differentially methylated probes were identified by fitting the DNA methylation data to a linear model. As the proportion of males and females in the samples of at least one of the groups was skewed, sex was included as a co-variate in the model, as was the proportion of memory CD4+ T cells in order to account for possible differences in T cell subtypes. Moderated t-statistics, log2 Fold Change (logFC) and p-values were computed for each investigated probe. The logFC values represents the average beta methylation difference (from hereon referred to as mean methylation difference, MMD) between the respective active intervention groups in relation to the double placebo (PP) group. P-values were adjusted for multiple testing using the Benjamini-Hochberg procedure for False Discovery Rate (FDR) correction. Differentially methylated CpGs (DMCs) were defined as probes having an FDR-adjusted p-value of less than 0.1 along with an MMD of > 5%. Volcano plots were constructed to graphically illustrate the distribution of the DMCs using the base R environment. Identification of genes of interest was conducted by mapping the DMCs to the genome, resulting in differentially methylated genes (DMG). The DMGs were defined as genes containing at least one DMC, where hypomethylation was defined as all DMCs being hypomethylated and vice versa for the hypermethylated DMGs. Genes containing both hypo- and hypermethylated sites were considered having a mixed methylation pattern. A total of three differential methylation comparisons were made between each of the treatment groups compared to the double placebo group. The quality of these comparisons was ascertained using several approaches. First of all, the genomic inflation rate was investigated for the t-statistics of each of the three comparisons described above, as the large number of statistical comparisons made within each analysis may render a considerable number of false positive results. The genomic inflation and pertaining bias were estimated and QQ-plots plotting the differential DNA methylation data against a normal distribution were constructed using the R package BACON^60^(version 1.14.0). The estimated inflation values for each of the three comparisons was very close to one in all cases, indicating that there was no substantial genomic inflation in the comparisons (Figure S11), and hence no correction was needed. Pathway enrichment analyses were performed on the identified DMCs from the differential DNA methylation analyses, using the clusterProfiler package (version 3.14.3) and the KEGG PATHWAY database (release 94 2020/04). An adjusted p-value cut-off of 0.1 was chosen to narrow down the number of pathways.

### Network analyses

To further study the biological relevance of the findings from the differential methylation analyses, the DMCs identified in the different comparisons described above were annotated to their corresponding gene symbols (DMGs) with the Illumina 850K platform annotation reference. A total of 93 deprecated unique gene terms from the platform reference were corrected to the most updated nomenclature using the org.Hs.eg.db R package (version 3.10.0). Thereafter, the DMGs were used as seeds to construct the inputs for the disease module identification pipeline with MODifieR^40^ R package (version 0.1.3, available at https://gitlab.com/Gustafsson-lab/MODifieR). Four different module defining algorithms from the package were implemented (Clique SuM, DIAMOnD, MCODE and Module Discoverer), and default parameters for the methods were used in each case. High confidence human protein-protein interactions (PPI) were retrieved from the STRING database (version 11.0, confidence score > 0.7). The four MODifieR methods were run for each comparison, resulting in a set of gene lists corresponding to the identified modules. Non-biased consensus modules were created for each comparison, by extracting genes found by all four module inference methods. Network visualisation was conducted in Cytoscape (version 3.7.2). Pathway enrichment analyses were performed on the module specific genes as described above to elucidate the role of the different intervention combinations on the CD4+ T cells. Furthermore, to unravel subclusters within the modules, visual inspection of the networks along with pathway enrichment analyses in the clusterProfiler R package (version 3.14.3, KEGG database) were performed. Clusters with three or more nodes were included in these analyses. Pathway enrichment analyses were also performed for unique and shared genes within the networks. To illustrate overlaps between shared and unique genes in the comparisons, Venn diagrams were constructed using the VennDiagram R package (version 1.6.20). Furthermore, to illustrate how the common genes may differ in methylation status between the comparisons, river plots were constructed using the riverplot R package (version 0.6).

### Comparisons of differential methylation and network results to publicly available allergy data set

As we were interested in the relevance of our findings to allergic disease, but do not yet have any data available on this from the children of the PROOM-3 study, we compared the findings of our study with publicly available data on allergic disease. The chosen data set was downloaded from Gene Expression Omnibus (https://www.ncbi.nlm.nih.gov/geo/query/acc.cgi?acc=GSE50222) and consisted of Illumina 450K data from CD4+ T cells from 8 adult patients *vs*. 8 controls with seasonal allergic rhinitis with samples from both within and out of season (in total 32 samples).^61^ The data were pre-processed by removing probes located on the sex chromosomes, and filtering was perfomed by removing probes having more than 20% NAs and imputing remaining NAs within retained probes using the KNN method (ChAMP, version 2.12.1). Thereafter, the data were normalised using the BMIQ method and SVD analyses followed (Figure S12) as described earlier, showing that no correction was needed. Subsequent differential DNA methylation analyses were performed using the limma package (version 3.42.2), and sex as well as season of sample taking (within or outside of pollen season) were added as co-variates in the linear models. Upon this, a total of 13102 DMCs were found for the data set, which were then compared to DMCs from our differential DNA methylation analyses and DMCs extracted from the genes identified in the respective network modules. Enrichment of any DMCs in these comparisons was performed using the enrichment function from the R package bc3net (version 1.0.4), based on a one-sided Fisher’s exact test. As the data set originated from 450K data, the intersection of all 450K probes and all MethylationEPIC 850K probes excluding the sex chromosomes was used as background for the analysis, and only DMCs that were present in the background were included. Corresponding pathway enrichment analyses for genes overlapping between any of the differentially methylated DMCs or consensus module originating DMCs with the allergy data set were performed as described above.

## Supporting information

Supplementary Appendix (incl. Fig S1-12, Tables S3, S6-11)

## Data Availability

The datasets used and/or analysed in the presented work are available on ArrayExpress, ID: E-MTAB-10341.

https://www.ebi.ac.uk/arrayexpress/experiments/E-MTAB-10341/

## ABBREVIATIONS

BMIQ: beta mixture quantile
CBMC: cord blood mononuclear cell
CFU: colony forming unit
CpG: Cytosine-phosphate-Guanine
DMC: differentially methylated CpG
DMG: differentially methylated gene
FDR: false discovery rate
GW: gestational week
*L. reuteri*: *Lactobacillus reuteri*
LP: single-treated *L*.*reuteri* + placebo group
Lω: double-treated *L*.*reuteri* + ω-3 group
MDS: multidimensional scaling
MMD: mean methylation difference
PBMC: peripheral blood mononuclear cell
PP: double placebo group
PROOM-3: PRObiotics and OMega-3 study
Pω: single-treated ω-3 + placebo group
SVD: singular value decomposition
ω-3: omega-3 fatty acids

## DECLARATIONS

### Ethics approval and consent to participate

Parents or legal guardians provided informed consent on behalf of the children prior to inclusion in the study. Ethical permission for this study has been granted by the Regional Ethics Committee for Human Research in Linköping (Dnr 2011/45-31).

### Consent for publication

Not applicable

### Author’s contributions

M.C.J and K.D. designed the study. K.D. and L.N. were responsible for the clinical evaluation of the children. J.H. has taken part in the sample collection and performed the experimental work presented in this paper. J.E. provided expertise on flow cytometry analyses, and M.G. on the performance of the statistical and bioinformatic analyses. D.M.E., E.O. and J.H. performed the statistical and bioinformatic analyses. J.H. presented the findings. All authors interpreted and discussed the results. J.H. drafted the manuscript. All authors contributed to and approved the final draft for publication.

## Acknowledgments

Firstly, we would like to thank the children and mothers included in this study for their participation, and the personnel at the Allergy Center, Linköping for professionally taking care of the families throughout the study period, as well as handling samples in preparation for laboratory work. Our warmest thanks go to our skilled laboratory technicians Camilla Janefjord and Anne-Marie Fornander, for their unfailing collection of samples from the PROOM-3 study. We would also like to thank Simon Söderholm for valuable input in the initial analyses of the DNA methylation data. Lastly, we would like to acknowledge BioGaia^®^ AB and Orkla Health for sponsoring the dietary supplements used in this study, as well as the SNP&SEQ-technology platform at SciLifeLab Uppsala University for yet another constructive collaboration.

## Author’s information

Not applicable

