## Supplementary Appendix (incl. Fig S1-12, Tables S3, S6-11) for "Combined prenatal *Lactobacillus reuteri* and ω-3 supplementation synergistically modulates DNA methylation in neonatal T helper cells"

Including Figures S1-S12 and Tables S3, S6-S11. Tables S1-S2, S4-S5 are supplied in separate files.

**RESULTS – SUPPLEMENTARY FIGURES AND TABLES**

**Figure S1.**
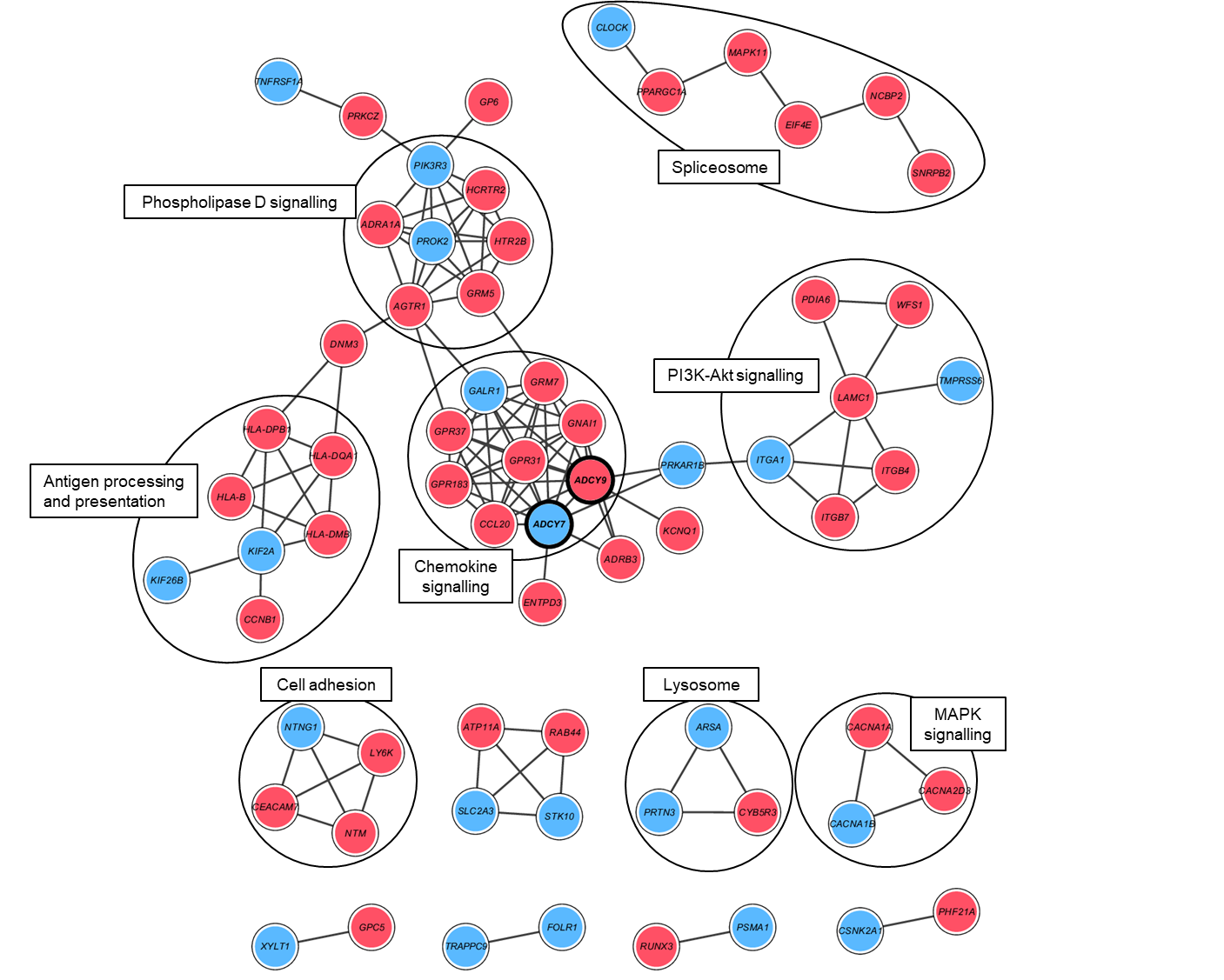

**LP *vs.* PP**

**Figure S1.** Visualisation of the consensus module created with the DMGs from the differential methylation comparison between the single treated *L.reuteri*/placebo (LP) and the double placebo (PP) group. Nodes represent genes and connecting lines represent protein-protein interactions (STRING combined score > 0.7) within the network. Red nodes illustrate hypermethylated genes, whereas blue nodes indicate hypomethylated genes. Black lines enclose biologically relevant clusters identified by bioinformatic pathway enrichment analyses. DMGs – differentially methylated genes, L – *Lactobacillus reuteri*, P – placebo.

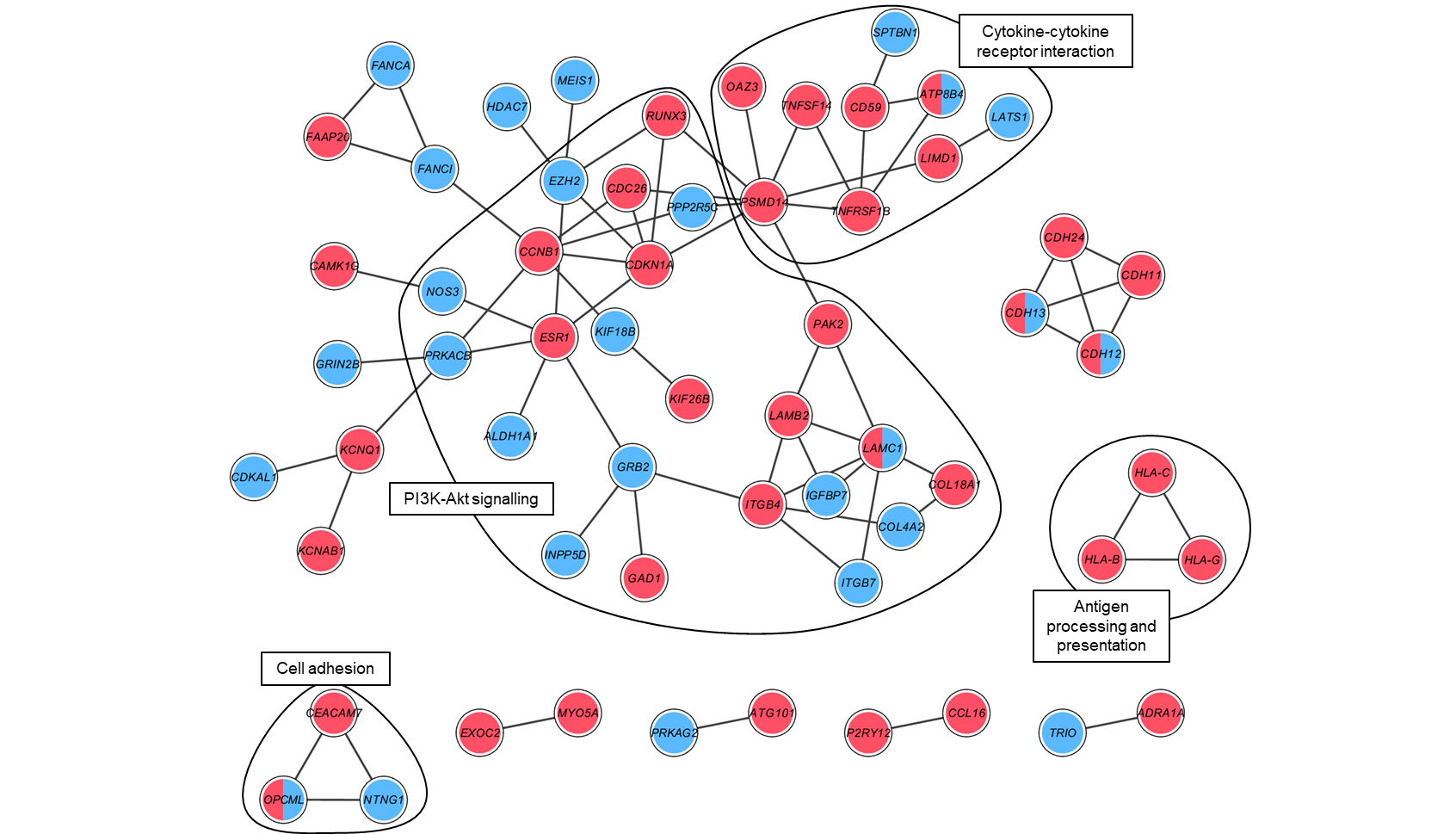
**Figure S2.**

**Pω *vs.* PP**

**Figure S2.** Visualisation of the consensus module created with the DMGs from the differential methylation comparison between the single treated placebo/ω-3 (Pω) and the double placebo (PP) group. Nodes represent genes and connecting lines represent protein-protein interactions (STRING combined score > 0.7) within the network. Red nodes illustrate hypermethylated genes, blue nodes indicate hypomethylated genes and both colours denote mixed methylation patterns. Black lines enclose biologically relevant clusters identified by bioinformatic pathway enrichment analyses. DMGs – differentially methylated genes, ω – ω-3 fatty acids, P – placebo.

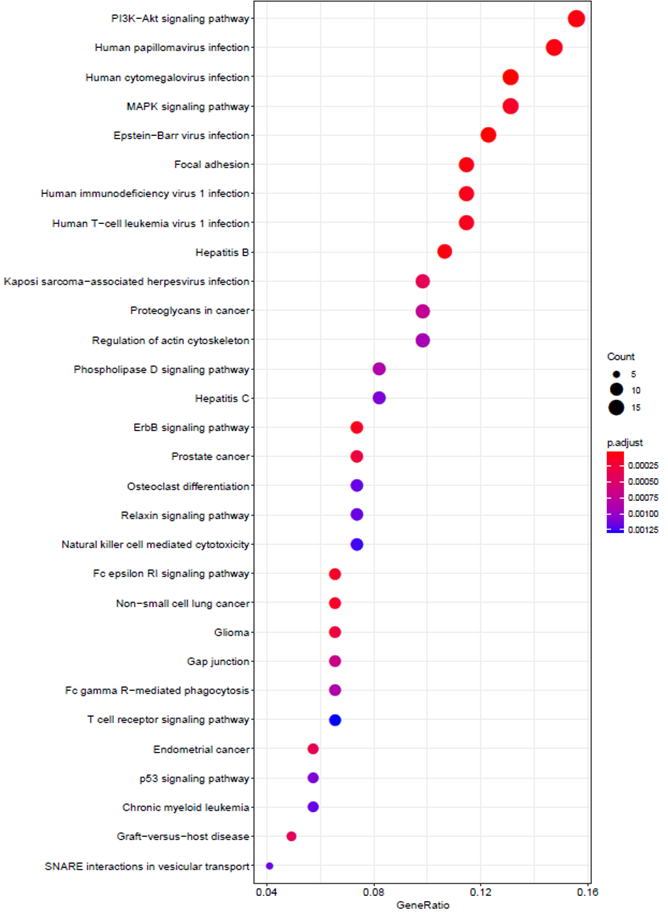
**Figure S3.**

Lω *vs.* PP

**Figure S3.** A dot plot illustrating the enriched pathways obtained by including all module genes from the comparison between the double treated *L.reuteri*/ω-3 (Lω) and the double placebo (PP) group. The x-axis represents the gene ratio, the dot size corresponds to the gene count, and the red-purple gradient dot colour represents adjusted enrichment P-values for the top 30 corresponding pathways designated on the left-hand side of the graph. L – *Lactobacillus reuteri*, ω – ω-3 fatty acids, P – placebo.

**Figure S4**

LP *vs.* PP

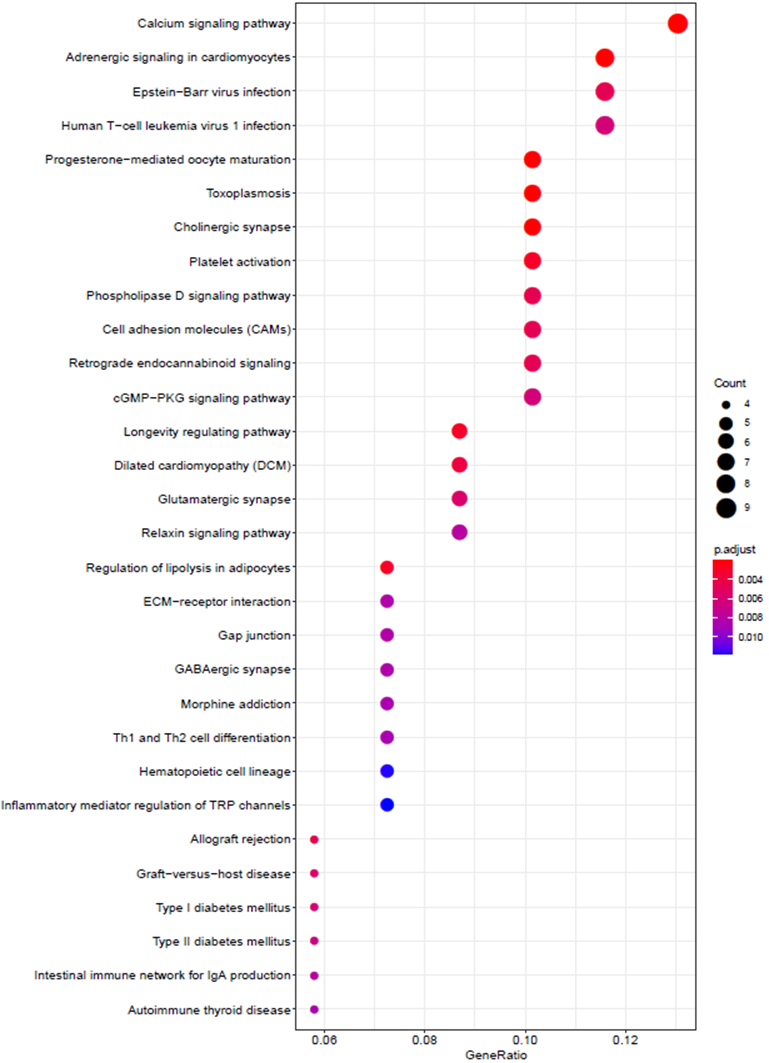
**Figure S4.** A dot plot illustrating the enriched pathways obtained by including all module genes from the comparison between the single treated *L.reuteri*/placebo (LP) and the double placebo (PP) group. The x-axis represents the gene ratio, the dot size corresponds to the gene count, and the red-purple gradient dot colour represents adjusted enrichment P-values for the top 30 corresponding pathways designated on the left-hand side of the graph. L – *Lactobacillus reuteri*, ω – ω-3 fatty acids, P – placebo.

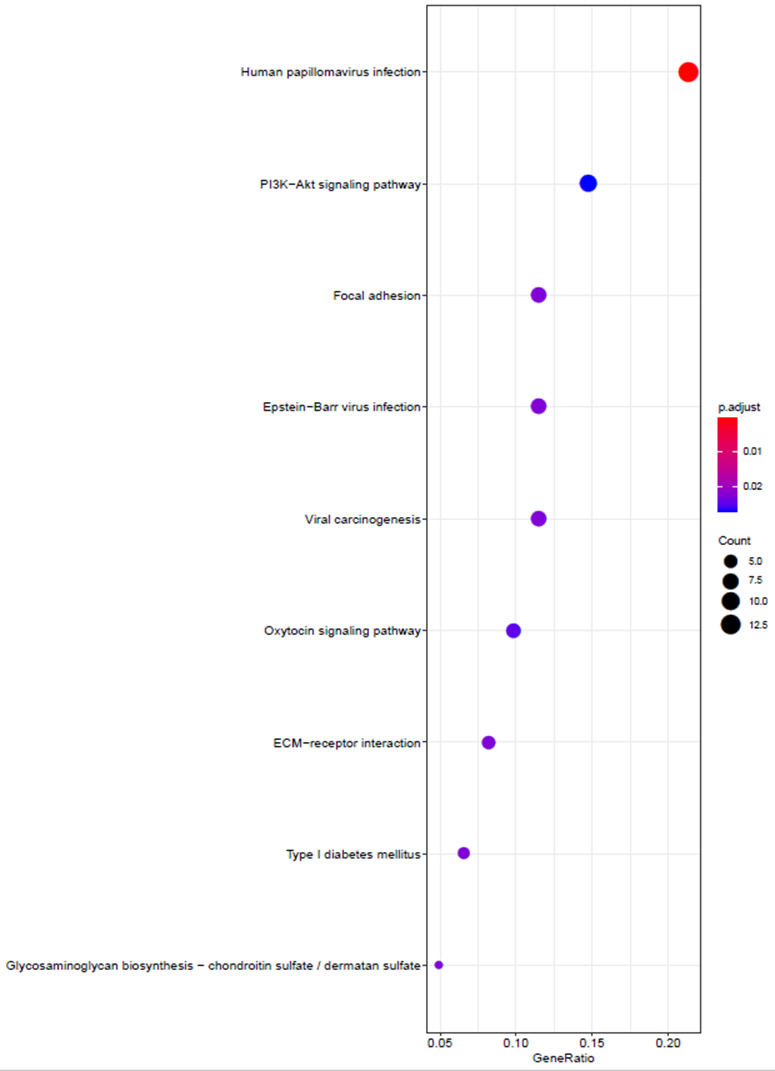
**Figure S5.**

Pω *vs.* PP

**Figure S5.** A dot plot illustrating the enriched pathways obtained by including all module genes from the comparison between the single treated placebo/ω-3 (Pω) and the double placebo (PP) group. The x-axis represents the gene ratio, the dot size corresponds to the gene count, and the red-purple gradient dot colour represents adjusted enrichment P-values for the corresponding pathways designated on the left-hand side of the graph. L – *Lactobacillus reuteri*, ω – ω-3 fatty acids, P – placebo.

**Figure S6.**

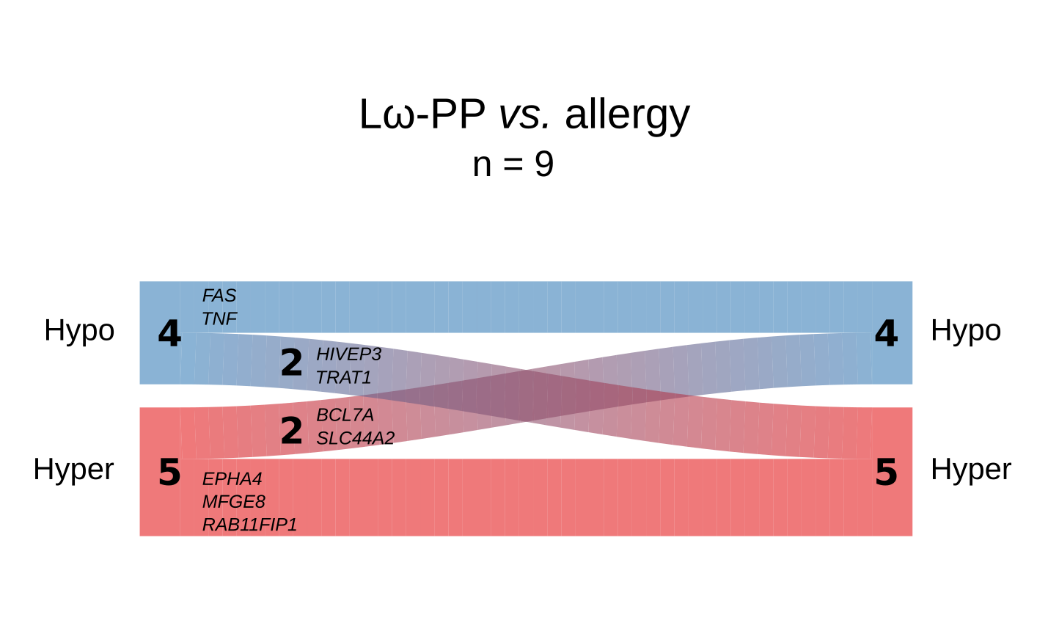

**Figure S6.** River plot illustrating the methylation status for overlapping genes being shared between the double-treated Lω group and the independent allergy data set. Genes are represented as flows of size proportional to number of genes within their methylation status in a particular set, and differences between sets are shown as connections between flows. The respective gene names are denoted next to each flow. Red represents hypermethylation and blue hypomethylation. The number of genes in the respective flows are denoted in the graph. L – Lactobacillus reuteri, ω – ω-3 fatty acids, P – placebo

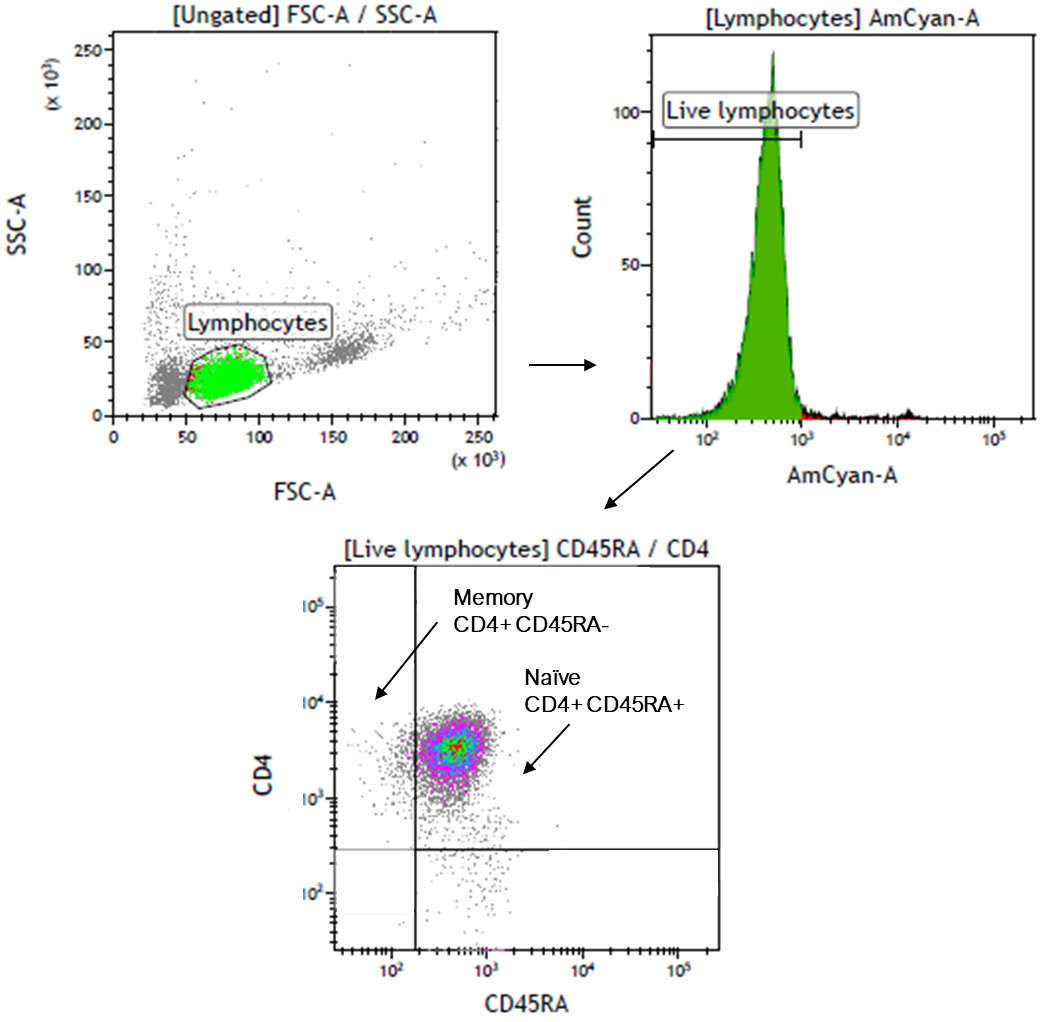
**Figure S7.**

**Figure S7.** Gating strategy for analysis of CD45RA+/- expression in CD4+ cells from cord blood mononuclear cells. Lymphocytes were defined using the FSC and SSC features. Thereafter, live cells within the lymphocyte population were determined using Aqua LIVE/DEAD (AmCyan-A) staining, and further gated to distinguish naïve (CD4+/CD45RA+) from memory (CD4+CD45RA-) cells. FSC – forward scatter, SSC – side scatter.

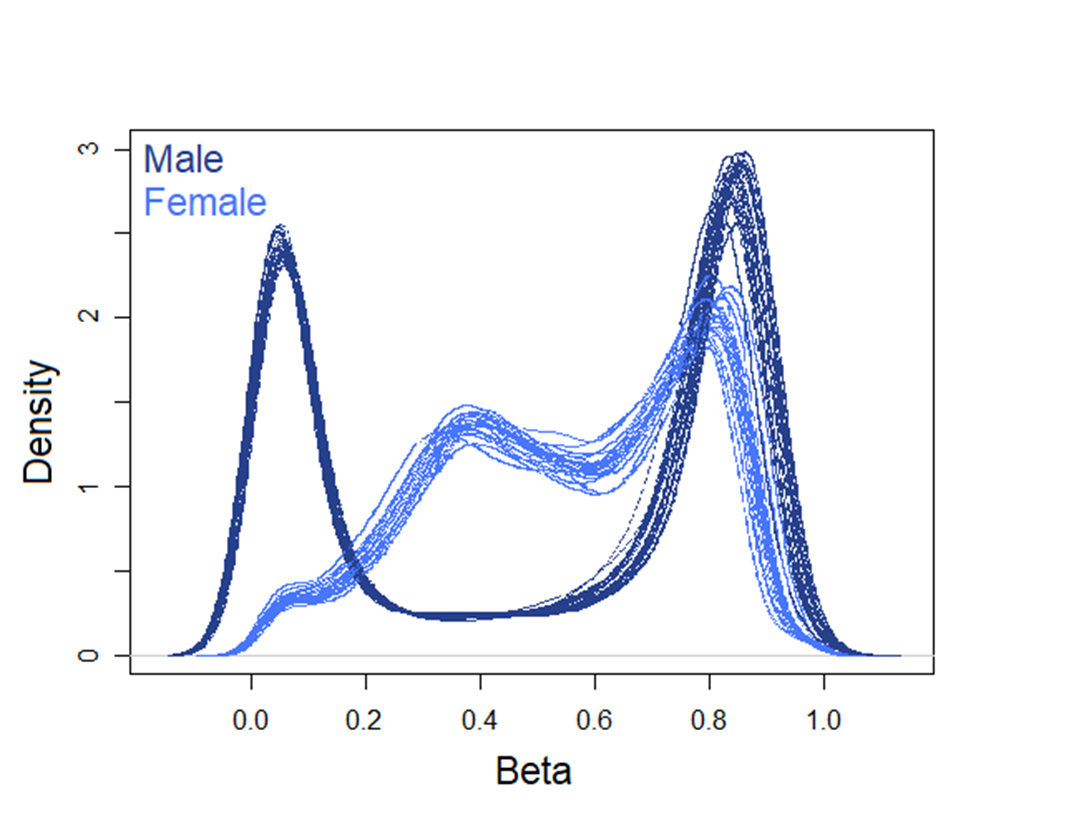
**Figure S8.**

**Figure S8.** Graphical representation of the density of the beta value distribution on the X chromosomes of males and females in raw DNA methylation data. Male samples are illustrated in dark blue, females in light blue.

**Figure S9.**

**b**

**a**

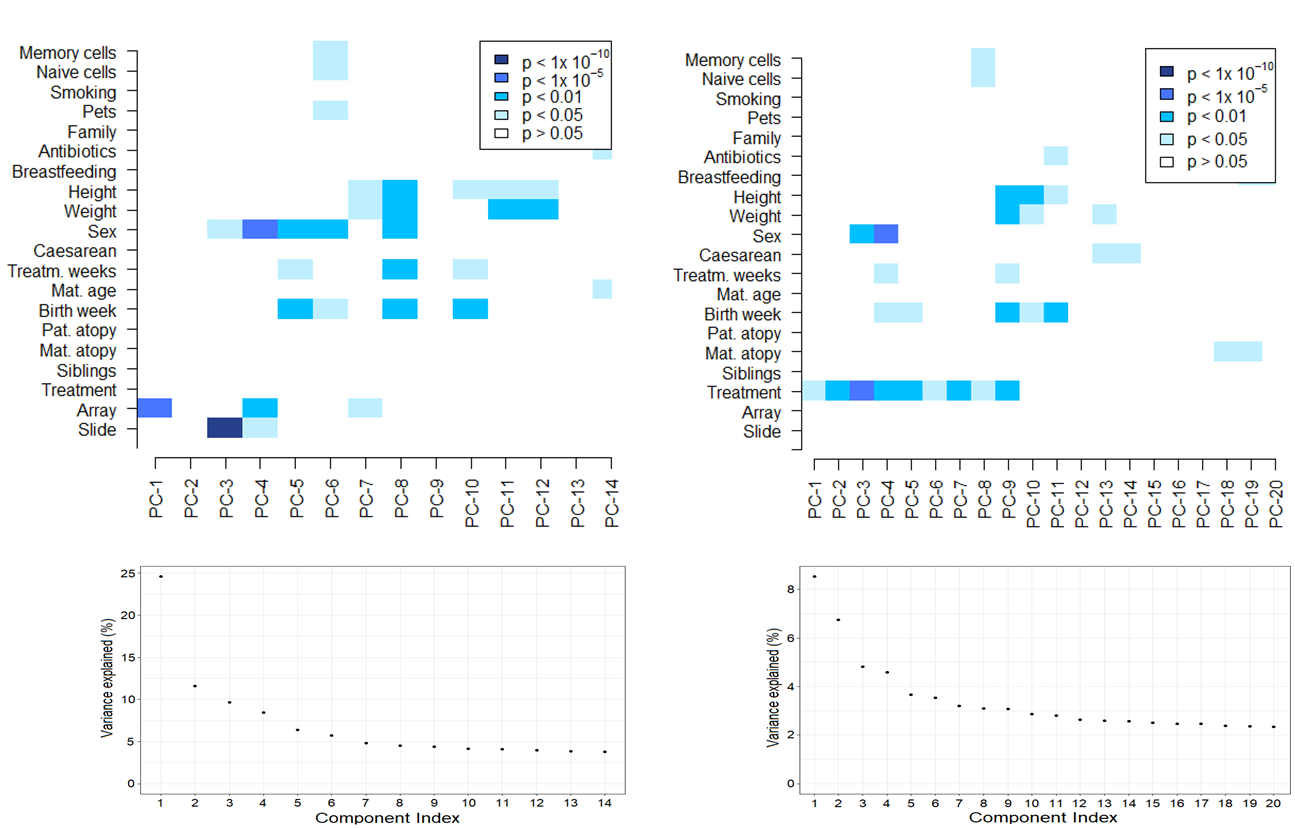

**Figure S9.** Graphical representations of Singular value decomposition (SVD) analyses for identification of major components of variation using the ChAMP package. Analyses on the filtered and BMIQ normalised data are displayed in panel a, and panel b shows the data upon correction for technical variables (array and slide). The lower panels present the % variance explained by each of the illustrated components in the SVD analyses. BMIQ – Beta Mixture Quantile.

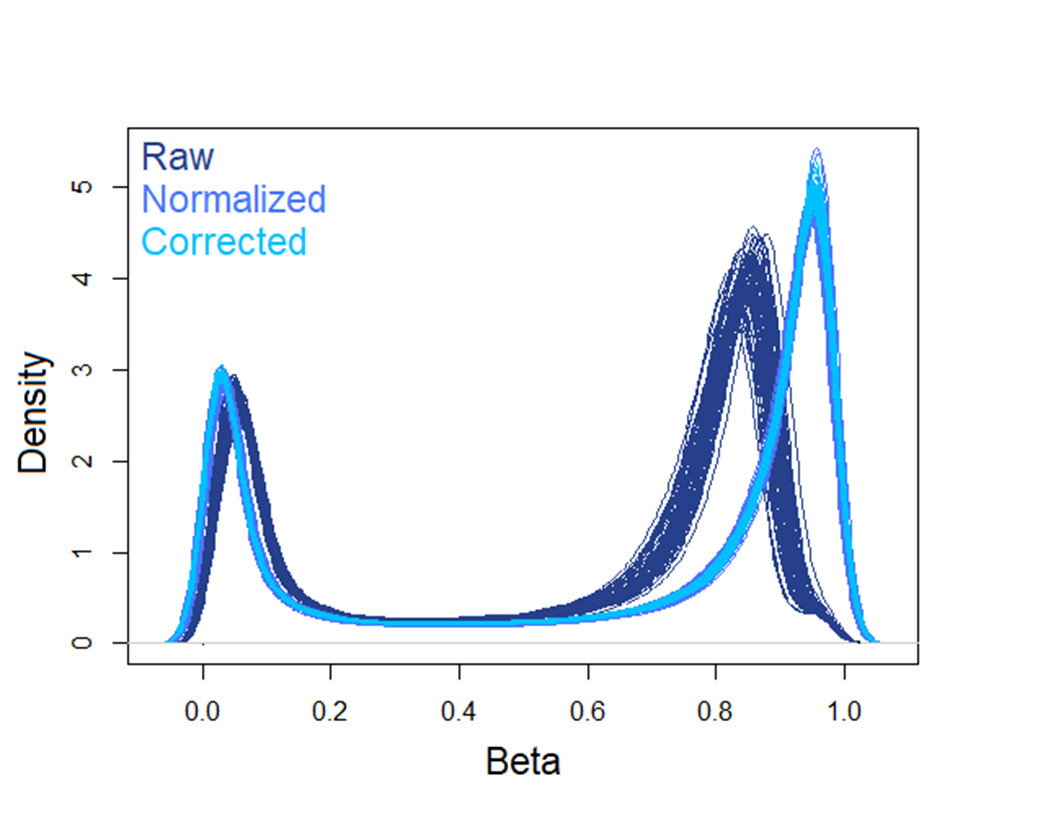
**Figure S10.**

**Figure S10.** Graphical representation of the density of the beta value distribution on the autosomal chromosomes prior to and after normalisation and correction. Raw DNA methylation data are depicted in dark blue, BMIQ-normalised data in light blue and batch corrected data (using the ChAMP package in both cases) are illustrated in coral blue.

**Figure S**
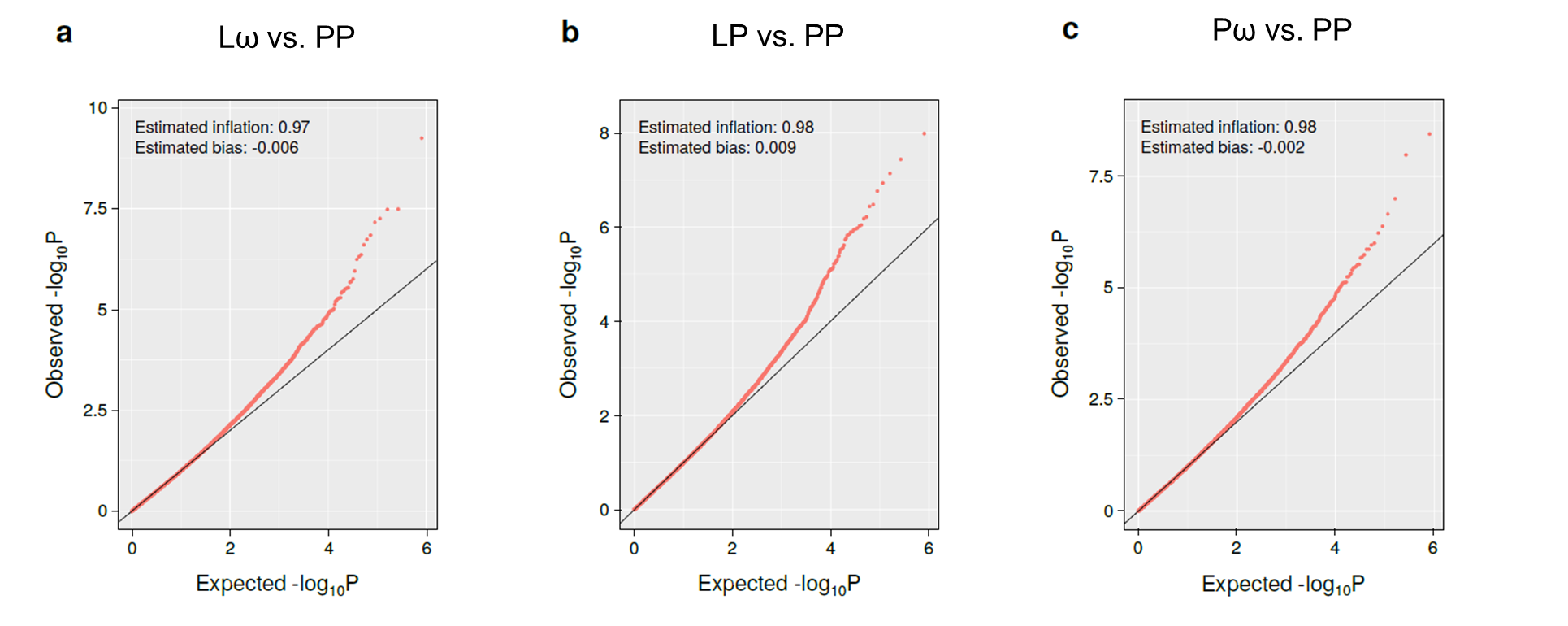
**11**

**Figure S11.** Plots illustrating the observed *vs.* expected –log10 of the obtained p-values from each of the three differential DNA methylation comparisons, with accompanying estimated values of genomic inflation and bias. Calculations were performed using the R package BACON. L – *Lactobacillus reuteri*, ω – ω-3 fatty acids, P – placebo.

**Figure S12.**

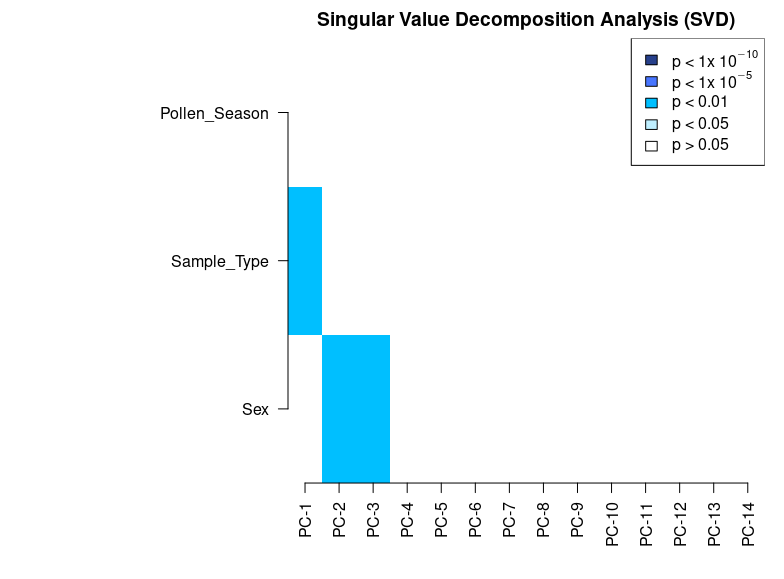

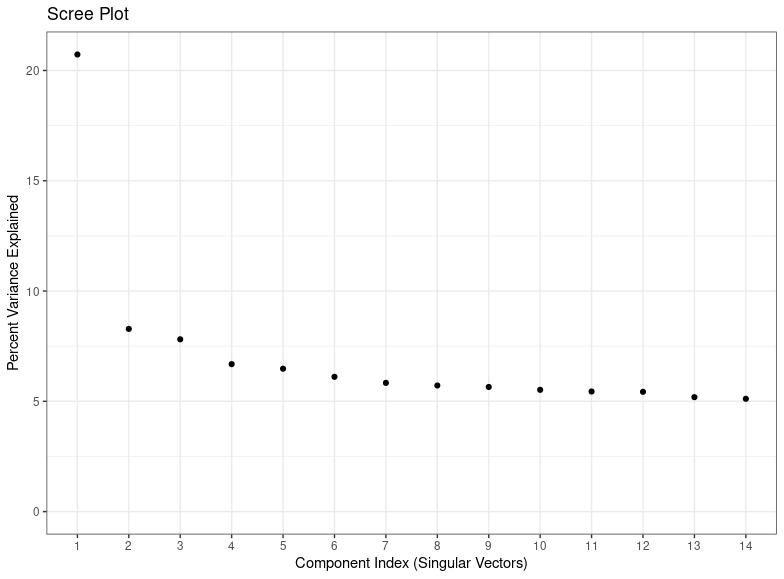

**Figure S12.** Graphical representation of Singular value decomposition (SVD) analyses for identification of major components of variation using the ChAMP package in the independent allergy data set. Analyses on the pre-processed, filtered and BMIQ normalised data are displayed in panel A, showing the major components of variation in the data. Panel B shows the Scree plot illustrating corresponding percent variance explained for each principal component. BMIQ – Beta Mixture Quantile.

**Table S1.** Table of extracted differentially methylated CpGs (DMCs) with corresponding mean methylation difference (MMD) from the three treatment comparisons (Lω, LP and Pω *vs.* PP, respectively). DMCs were defined as having a MMD of > ±5% along with an FDR-corrected p-value of <0.1. L – *Lactobacillus reuteri*, ω – ω-3 fatty acids, P – placebo. Supplied in a separate file.

**Table S2.** Table of differentially methylated genes (DMGs) with corresponding direction of methylation from the three treatment comparisons (Lω, LP and Pω). DMGs were defined as genes with at least one DMC, and the direction of methylation depended on the direction of methylation for all DMCs in the gene. Hypomethylation is defined as > -5% mean methylation difference (MMD), hypermethylation as > +5% MMD and mixed methylation genes contained DMCs of both directions. L – *Lactobacillus reuteri*, ω – ω-3 fatty acids, P – placebo. Supplied in a separate file.

**Table S4.** Table of pathways obtained from pathway enrichment analyses on DMCs from the Lω *vs.* PP comparison. Analyses were conducted using clusterProfiler R package and the KEGG PATHWAY database. Pathways with an adjusted p-value of 0.1 or lower were considered significantly relevant.

**Table S5.** Table of pathways obtained from pathway enrichment analyses on DMCs from the LP *vs.* PP comparison. Analyses were conducted using clusterProfiler R package and the KEGG PATHWAY database. Pathways with an adjusted p-value of 0.1 or lower were considered significantly relevant.

**Table S3.** Table of differentially methylated genes that were shared between the three treatment groups (Lω, LP and Pω). L – *Lactobacillus reuteri*, ω – ω-3 fatty acids, P – placebo.

| Shared differentially  methylated genes  (n=72) | |
| --- | --- |
| *FCRLB* | *SLC45A4* |
| *PTPRN2* | *NBL1* |
| *RPS6KA2* | *ZFR* |
| *RFPL2* | *HIVEP2* |
| *SYT17* | *NAV2* |
| *UNC45A* | *CSGALNACT1* |
| *ZSCAN12P1* | *KDM5B* |
| *PHF21A* | *B3GNT1* |
| *KIF26B* | *KCNG2* |
| *C1QL3* | *SCGB1A1* |
| *C17orf54* | *ZNF253* |
| *KRTAP12-3* | *MYOF* |
| *MAGI2* | *CEACAM7* |
| *CRISPLD2* | *CCDC162P* |
| *ZSCAN12L1* | *GRIK4* |
| *WNK4* | *MCTP1* |
| *SLC1A7* | *SYNPO2* |
| *NTNG1* | *OTX2-AS1* |
| *AGPAT3* | *IQGAP1* |
| *SLC35F3* | *REM2* |
| *PKNOX2* | *SORCS2* |
| *MICAL3* | *ITGB7* |
| *LGR6* | *RFTN1* |
| *OLFM1* | *DEFB115* |
| *C6orf123* | *ABI1* |
| *MIR548N* | *DGKG* |
| *CDH13* | *RUNX3* |
| *MX2* | *PLA2G4C* |
| *ZNF710* | *LOC101929241* |
| *LAMC1* | *CDC42BPA* |
| *FAM38A* | *NPHP1* |
| *CRTAC1* | *SORCS1* |
| *MIB2* | *MRGPRF* |
| *S100P* | *CRX* |
| *CD164* | *HLA-B* |
| *C10orf93* | *GP6* |

**Table S6.** A table of module genes from the Lω comparison, displaying Entrez-ID, direction of methylation (hypo-, hyper- or mixed methylation), the number of connecting genes within network module and absolute mean methylation difference. L – *Lactobacillus reuteri*, MMD – mean methylation difference, ω – ω-3 fatty acids, P – placebo.

| Module genes from the Lω comparison | | | | |
| --- | --- | --- | --- | --- |
| Gene symbol | **Entrez** | **Direction of  methylation** | **Degree of  connectivity** | **Absolute MMD** |
| *FYN* | 2534 | Hyper | 16 | 0,071 |
| *GNB5* | 10681 | Hyper | 14 | 0,082 |
| *SNAP23* | 8773 | Hyper | 13 | 0,055 |
| *LAMC1* | 3915 | Hyper | 12 | 0,095 |
| *IL6* | 3569 | Hypo | 10 | 0,051 |
| *RAF1* | 5894 | Hyper | 10 | 0,055 |
| *ADCY9* | 115 | Mixed | 9 | 0,074 |
| *CUL1* | 8454 | Hyper | 9 | 0,056 |
| *TNF* | 7124 | Hypo | 9 | 0,053 |
| *AP1M1* | 8907 | Hyper | 8 | 0,063 |
| *CREB1* | 1385 | Hypo | 8 | 0,052 |
| *GRB2* | 2885 | Hyper | 8 | 0,059 |
| *HMOX2* | 3163 | Hyper | 8 | 0,059 |
| *LPAR1* | 1902 | Hyper | 8 | 0,074 |
| *PAK1* | 5058 | Hyper | 8 | 0,053 |
| *PRKACB* | 5567 | Hypo | 8 | 0,055 |
| *SEC22B* | 9554 | Hypo | 8 | 0,058 |
| *CD47* | 961 | Hypo | 7 | 0,068 |
| *CDC23* | 8697 | Hypo | 7 | 0,060 |
| *ITGB5* | 3693 | Hyper | 7 | 0,079 |
| *KCNAB2* | 8514 | Hyper | 7 | 0,090 |
| *MAP2K7* | 5609 | Hyper | 7 | 0,057 |
| *PAK2* | 5062 | Hyper | 7 | 0,060 |
| *PRTN3* | 5657 | Hypo | 7 | 0,176 |
| *SDCBP* | 6386 | Hypo | 7 | 0,052 |
| *SLC2A3* | 6515 | Hypo | 7 | 0,069 |
| *SLC44A2* | 57153 | Hyper | 7 | 0,051 |
| *STK10* | 6793 | Hypo | 7 | 0,086 |
| *ADA2* | 51816 | Hypo | 6 | 0,062 |
| *AKT3* | 10000 | Hyper | 6 | 0,051 |
| *CDKN1A* | 1026 | Hyper | 6 | 0,095 |
| *CPPED1* | 55313 | Hyper | 6 | 0,052 |
| *CYB5R3* | 1727 | Hyper | 6 | 0,061 |
| *HTR2B* | 3357 | Hyper | 6 | 0,054 |
| *IST1* | 9798 | Hyper | 6 | 0,052 |
| *MAN2B1* | 4125 | Hypo | 6 | 0,063 |
| *MFGE8* | 4240 | Hyper | 6 | 0,104 |
| *PRKCA* | 5578 | Hyper | 6 | 0,066 |
| *SYK* | 6850 | Hyper | 6 | 0,055 |
| *ADRB3* | 155 | Hyper | 5 | 0,089 |
| *ATM* | 472 | Hypo | 5 | 0,057 |
| *CALCR* | 799 | Hypo | 5 | 0,053 |
| *EPHB3* | 2049 | Hyper | 5 | 0,062 |
| *FAM20C* | 56975 | Hyper | 5 | 0,069 |
| *FAS* | 355 | Hypo | 5 | 0,059 |
| *FSHR* | 2492 | Hyper | 5 | 0,051 |
| *GPR27* | 2850 | Hyper | 5 | 0,138 |
| *ITGA8* | 8516 | Hypo | 5 | 0,062 |
| *ITGB7* | 3695 | Hypo | 5 | 0,051 |
| *LTB4R2* | 56413 | Hyper | 5 | 0,072 |
| *NPFFR2* | 10886 | Hypo | 5 | 0,053 |
| *PDIA6* | 10130 | Hyper | 5 | 0,079 |
| *PROK2* | 60675 | Hyper | 5 | 0,108 |
| *RCHY1* | 25898 | Hyper | 5 | 0,058 |
| *RING1* | 6015 | Hyper | 5 | 0,053 |
| *UBE2I* | 7329 | Hyper | 5 | 0,061 |
| *WFS1* | 7466 | Hyper | 5 | 0,052 |
| *AGRN* | 375790 | Hypo | 4 | 0,059 |
| *ARFGAP3* | 26286 | Hyper | 4 | 0,052 |
| *ASB10* | 136371 | Hyper | 4 | 0,054 |
| *CCL16* | 6360 | Hyper | 4 | 0,069 |
| *CEACAM7* | 1087 | Hyper | 4 | 0,068 |
| *ERBB4* | 2066 | Hyper | 4 | 0,054 |
| *FBXL7* | 23194 | Hyper | 4 | 0,056 |
| *GPR31* | 2853 | Hyper | 4 | 0,056 |
| *HLA-DRB1* | 3123 | Hyper | 4 | 0,222 |
| *KIF16B* | 55614 | Hyper | 4 | 0,076 |
| *KIF25* | 3834 | Hypo | 4 | 0,063 |
| *KIF26B* | 55083 | Hyper | 4 | 0,063 |
| *MDGA1* | 266727 | Hyper | 4 | 0,109 |
| *NTNG1* | 22854 | Hypo | 4 | 0,074 |
| *OPCML* | 4978 | Hypo | 4 | 0,132 |
| *PGR* | 5241 | Hyper | 4 | 0,050 |
| *PSCA* | 8000 | Hyper | 4 | 0,052 |
| *RB1* | 5925 | Hyper | 4 | 0,100 |
| *RUNX1* | 861 | Hypo | 4 | 0,054 |
| *TP73* | 7161 | Hyper | 4 | 0,056 |
| *ACTN4* | 81 | Hyper | 3 | 0,055 |
| *AXIN1* | 8312 | Hyper | 3 | 0,080 |
| *CASP9* | 842 | Hypo | 3 | 0,065 |
| *EED* | 8726 | Hypo | 3 | 0,067 |
| *ERCC4* | 2072 | Hyper | 3 | 0,051 |
| *HLA-B* | 3106 | Hyper | 3 | 0,262 |
| *ITGAE* | 3682 | Hyper | 3 | 0,059 |
| *MRPS2* | 51116 | Hyper | 3 | 0,054 |
| *MUC17* | 140453 | Hyper | 3 | 0,055 |
| *RPS25* | 6230 | Hyper | 3 | 0,051 |
| *AKAP9* | 10142 | Hyper | 2 | 0,062 |
| *ATG5* | 9474 | Hypo | 2 | 0,053 |
| *BET1L* | 51272 | Hyper | 2 | 0,069 |
| *CRNKL1* | 51340 | Hyper | 2 | 0,056 |
| *DAB1* | 1600 | Hyper | 2 | 0,066 |
| *DDB2* | 1643 | Hyper | 2 | 0,052 |
| *EPHA4* | 2043 | Hyper | 2 | 0,092 |
| *EPS15L1* | 58513 | Hyper | 2 | 0,076 |
| *GLI2* | 2736 | Hyper | 2 | 0,060 |
| *GP6* | 51206 | Hyper | 2 | 0,060 |
| *HLA-DOB* | 3112 | Hyper | 2 | 0,058 |
| *HNRNPM* | 4670 | Hyper | 2 | 0,083 |
| *HTR3A* | 3359 | Hypo | 2 | 0,086 |
| *LDLRAP1* | 26119 | Hyper | 2 | 0,057 |
| *MRPL28* | 10573 | Hyper | 2 | 0,157 |
| *MRPL51* | 51258 | Hyper | 2 | 0,065 |
| *PDGFRA* | 5156 | Hyper | 2 | 0,052 |
| *PHF21A* | 51317 | Hyper | 2 | 0,067 |
| *RELB* | 5971 | Hyper | 2 | 0,104 |
| *RHOH* | 399 | Hyper | 2 | 0,110 |
| *RHOJ* | 57381 | Hyper | 2 | 0,055 |
| *RHOT1* | 55288 | Hyper | 2 | 0,064 |
| *RUNX3* | 864 | Hyper | 2 | 0,163 |
| *SALL4* | 57167 | Hyper | 2 | 0,063 |
| *SGIP1* | 84251 | Hypo | 2 | 0,069 |
| *STX19* | 415117 | Hypo | 2 | 0,051 |
| *STX2* | 2054 | Hypo | 2 | 0,061 |
| *TRPC6* | 7225 | Hyper | 2 | 0,077 |
| *WDR33* | 55339 | Hyper | 2 | 0,113 |
| *AGO2* | 27161 | Hyper | 1 | 0,054 |
| *ASAP1* | 50807 | Hyper | 1 | 0,052 |
| *ASAP2* | 8853 | Hyper | 1 | 0,050 |
| *ATG101* | 60673 | Hyper | 1 | 0,076 |
| *ATG4C* | 84938 | Hyper | 1 | 0,052 |
| *BCL7A* | 605 | Hyper | 1 | 0,058 |
| *BRD7* | 29117 | Mixed | 1 | 0,072 |
| *CACNA1A* | 773 | Hyper | 1 | 0,066 |
| *CDH12* | 1010 | Hypo | 1 | 0,073 |
| *CDH13* | 1012 | Hyper | 1 | 0,087 |
| *CHSY1* | 22856 | Hypo | 1 | 0,093 |
| *CLOCK* | 9575 | Hypo | 1 | 0,055 |
| *CSGALNACT1* | 55790 | Hypo | 1 | 0,091 |
| *EEF1A2* | 1917 | Hyper | 1 | 0,054 |
| *EIF3E* | 3646 | Hypo | 1 | 0,068 |
| *ELL* | 8178 | Hyper | 1 | 0,081 |
| *FGFR2* | 2263 | Hyper | 1 | 0,174 |
| *GALNT13* | 114805 | Mixed | 1 | 0,055 |
| *GALNT7* | 51809 | Hyper | 1 | 0,053 |
| *GALNT9* | 50614 | Hypo | 1 | 0,060 |
| *HOXB6* | 3216 | Hypo | 1 | 0,073 |
| *HOXC4* | 3221 | Hyper | 1 | 0,068 |
| *IDI1* | 3422 | Hypo | 1 | 0,075 |
| *INSIG1* | 3638 | Hyper | 1 | 0,072 |
| *MAP4K3* | 8491 | Hypo | 1 | 0,058 |
| *MEIS1* | 4211 | Hypo | 1 | 0,058 |
| *MYO5A* | 4644 | Hyper | 1 | 0,060 |
| *NPAS2* | 4862 | Hyper | 1 | 0,061 |
| *NSUN2* | 54888 | Hypo | 1 | 0,079 |
| *POLR1B* | 84172 | Hyper | 1 | 0,055 |
| *PPP2R2C* | 5522 | Hypo | 1 | 0,065 |
| *SCNN1A* | 6337 | Hyper | 1 | 0,054 |
| *TANK* | 10010 | Hypo | 1 | 0,071 |
| *TMPRSS6* | 164656 | Hypo | 1 | 0,062 |
| *TNIK* | 23043 | Hypo | 1 | 0,061 |
| *TRAT1* | 50852 | Hypo | 1 | 0,051 |
| *WNT3A* | 89780 | Hyper | 1 | 0,062 |
| *ALDH1A1* | 216 | Hypo | 0 | 0,050 |
| *C9orf47* | 286223 | Hyper | 0 | 0,074 |
| *DSE* | 29940 | Hyper | 0 | 0,059 |
| *EXOC6B* | 23233 | Hypo | 0 | 0,063 |
| *HIVEP3* | 59269 | Hypo | 0 | 0,050 |
| *INPP5A* | 3632 | Hyper | 0 | 0,164 |
| *LOR* | 4014 | Hypo | 0 | 0,097 |
| *LOX* | 4015 | Hypo | 0 | 0,097 |
| *LTF* | 4057 | Hyper | 0 | 0,062 |
| *MKLN1* | 4289 | Hypo | 0 | 0,053 |
| *OS9* | 10956 | Hyper | 0 | 0,060 |
| *PIP5K1B* | 8395 | Hyper | 0 | 0,060 |
| *PRICKLE1* | 144165 | Hyper | 0 | 0,063 |
| *RAB11FIP1* | 80223 | Hyper | 0 | 0,077 |
| *RARRES2* | 5919 | Hyper | 0 | 0,054 |
| *RTN4* | 57142 | Hyper | 0 | 0,057 |
| *SKA2* | 348235 | Hyper | 0 | 0,068 |

**Table S7.** A table of module genes from the LP comparison, displaying Entrez-ID, direction of methylation (hypo-, hyper- or mixed methylation), the number of connecting genes within network module and absolute mean methylation difference. L – *Lactobacillus reuteri*, MMD – mean methylation difference, P – placebo.

| Module genes from the LP comparison | | | | |
| --- | --- | --- | --- | --- |
| Gene symbol | **Entrez** | **Direction of  methylation** | **Degree of  connectivity** | **Absolute MMD** |
| *ADCY7* | 113 | Hypo | 11 | 0,079 |
| *ADCY9* | 115 | Hyper | 11 | 0,058 |
| *AGTR1* | 185 | Hyper | 9 | 0,054 |
| *GALR1* | 2587 | Hypo | 9 | 0,069 |
| *GNAI1* | 2770 | Hyper | 9 | 0,080 |
| *GPR37* | 2861 | Hyper | 9 | 0,053 |
| *GRM7* | 2917 | Hyper | 9 | 0,058 |
| *CCL20* | 6364 | Hyper | 8 | 0,067 |
| *GPR183* | 1880 | Hyper | 8 | 0,051 |
| *GPR31* | 2853 | Hyper | 8 | 0,059 |
| *PIK3R3* | 8503 | Hypo | 8 | 0,051 |
| *GRM5* | 2915 | Hyper | 7 | 0,061 |
| *ADRA1A* | 148 | Hyper | 6 | 0,052 |
| *HCRTR2* | 3062 | Hyper | 6 | 0,065 |
| *HTR2B* | 3357 | Hyper | 6 | 0,074 |
| *LAMC1* | 3915 | Hyper | 6 | 0,063 |
| *PROK2* | 60675 | Hypo | 6 | 0,052 |
| *HLA-DPB1* | 3115 | Hyper | 5 | 0,062 |
| *HLA-DQA1* | 3117 | Hyper | 5 | 0,359 |
| *KIF2A* | 3796 | Hypo | 5 | 0,052 |
| *HLA-DMB* | 3109 | Hyper | 4 | 0,064 |
| *ITGA1* | 3672 | Hypo | 4 | 0,053 |
| *ADRB3* | 155 | Hyper | 3 | 0,100 |
| *ATP11A* | 23250 | Hyper | 3 | 0,057 |
| *CEACAM7* | 1087 | Hyper | 3 | 0,063 |
| *DNM3* | 26052 | Hyper | 3 | 0,056 |
| *HLA-B* | 3106 | Hyper | 3 | 0,064 |
| *ITGB4* | 3691 | Hyper | 3 | 0,055 |
| *ITGB7* | 3695 | Hyper | 3 | 0,057 |
| *LY6K* | 54742 | Hyper | 3 | 0,052 |
| *NTM* | 50863 | Hyper | 3 | 0,053 |
| *NTNG1* | 22854 | Hypo | 3 | 0,063 |
| *PRKAR1B* | 5575 | Hypo | 3 | 0,061 |
| *RAB44* | 401258 | Hyper | 3 | 0,060 |
| *SLC2A3* | 6515 | Hypo | 3 | 0,068 |
| *STK10* | 6793 | Hypo | 3 | 0,065 |
| *ARSA* | 410 | Hypo | 2 | 0,058 |
| *CACNA1A* | 773 | Hyper | 2 | 0,080 |
| *CACNA1B* | 774 | Hypo | 2 | 0,051 |
| *CACNA2D3* | 55799 | Hyper | 2 | 0,053 |
| *CYB5R3* | 1727 | Hyper | 2 | 0,069 |
| *EIF4E* | 1977 | Hyper | 2 | 0,077 |
| *MAPK11* | 5600 | Hyper | 2 | 0,052 |
| *NCBP2* | 22916 | Hyper | 2 | 0,054 |
| *PDIA6* | 10130 | Hyper | 2 | 0,057 |
| *PPARGC1A* | 10891 | Hyper | 2 | 0,052 |
| *PRKCZ* | 5590 | Hyper | 2 | 0,071 |
| *PRTN3* | 5657 | Hypo | 2 | 0,129 |
| *WFS1* | 7466 | Hyper | 2 | 0,051 |
| *CCNB1* | 891 | Hyper | 1 | 0,068 |
| *CLOCK* | 9575 | Hypo | 1 | 0,059 |
| *CSNK2A1* | 1457 | Hypo | 1 | 0,067 |
| *ENTPD3* | 956 | Hyper | 1 | 0,058 |
| *FOLR1* | 2348 | Hypo | 1 | 0,051 |
| *GP6* | 51206 | Hyper | 1 | 0,066 |
| *GPC5* | 2262 | Hyper | 1 | 0,064 |
| *KCNQ1* | 3784 | Hyper | 1 | 0,057 |
| *KIF26B* | 55083 | Hypo | 1 | 0,069 |
| *PHF21A* | 51317 | Hyper | 1 | 0,073 |
| *PSMA1* | 5682 | Hypo | 1 | 0,062 |
| *RUNX3* | 864 | Hyper | 1 | 0,128 |
| *SNRPB2* | 6629 | Hyper | 1 | 0,051 |
| *TMPRSS6* | 164656 | Hypo | 1 | 0,067 |
| *TNFRSF1A* | 7132 | Hypo | 1 | 0,068 |
| *TRAPPC9* | 83696 | Hypo | 1 | 0,127 |
| *XYLT1* | 64131 | Hypo | 1 | 0,052 |
| *ADAM22* | 53616 | Hyper | 0 | 0,059 |
| *ASAP1* | 50807 | Hyper | 0 | 0,050 |
| *C2CD5* | 9847 | Hypo | 0 | 0,064 |
| *CBFA2T3* | 863 | Hypo | 0 | 0,060 |
| *CD1A* | 909 | Hypo | 0 | 0,076 |
| *CDH13* | 1012 | Hyper | 0 | 0,091 |
| *CLDN15* | 24146 | Hypo | 0 | 0,061 |
| *CPS1* | 1373 | Hyper | 0 | 0,054 |
| *CSGALNACT1* | 55790 | Mixed | 0 | 0,133 |
| *CTTNBP2* | 83992 | Hyper | 0 | 0,066 |
| *CUX1* | 1523 | Hyper | 0 | 0,105 |
| *DSC3* | 1825 | Hyper | 0 | 0,068 |
| *GALNT7* | 51809 | Hyper | 0 | 0,099 |
| *GALNT9* | 50614 | Hyper | 0 | 0,051 |
| *GALNTL6* | 442117 | Hypo | 0 | 0,052 |
| *HOXA5* | 3202 | Hypo | 0 | 0,061 |
| *IDO1* | 3620 | Hyper | 0 | 0,051 |
| *INSIG1* | 3638 | Hyper | 0 | 0,060 |
| *KDM2B* | 84678 | Hypo | 0 | 0,058 |
| *KRT84* | 3890 | Hypo | 0 | 0,085 |
| *MKLN1* | 4289 | Hyper | 0 | 0,060 |
| *MRPL48* | 51642 | Hyper | 0 | 0,066 |
| *PARP1* | 142 | Hypo | 0 | 0,067 |
| *PRSS3* | 5646 | Hyper | 0 | 0,057 |
| *RAD51B* | 5890 | Hyper | 0 | 0,070 |
| *RB1CC1* | 9821 | Hypo | 0 | 0,056 |
| *RCHY1* | 25898 | Hyper | 0 | 0,051 |
| *RHOH* | 399 | Hyper | 0 | 0,098 |
| *ROR2* | 4920 | Hyper | 0 | 0,080 |
| *RTN4* | 57142 | Hypo | 0 | 0,055 |
| *THEMIS* | 387357 | Hyper | 0 | 0,051 |
| *TRPC6* | 7225 | Hyper | 0 | 0,074 |
| *USH2A* | 7399 | Hyper | 0 | 0,053 |
| *WWTR1* | 25937 | Hyper | 0 | 0,060 |

**Table S8.** A table of module genes from the Pω comparison, displaying Entrez-ID, direction of methylation (hypo-, hyper- or mixed methylation), the number of connecting genes within network module and absolute mean methylation difference. ω – ω-3 fatty acids, MMD – mean methylation difference, P – placebo.

| Module genes from the Pω comparison | | | | |
| --- | --- | --- | --- | --- |
| Gene symbol | **Entrez** | **Direction of  methylation** | **Degree of  connectivity** | **Absolute MMD** |
| *PSMD14* | 10213 | Hyper | 9 | 0,052 |
| *CCNB1* | 891 | Hyper | 6 | 0,057 |
| *CDKN1A* | 1026 | Hyper | 6 | 0,134 |
| *ESR1* | 2099 | Hyper | 6 | 0,057 |
| *LAMC1* | 3915 | Mixed | 6 | 0,089 |
| *EZH2* | 2146 | Hypo | 5 | 0,056 |
| *ITGB4* | 3691 | Hyper | 5 | 0,053 |
| *GRB2* | 2885 | Hypo | 4 | 0,053 |
| *LAMB2* | 3913 | Hyper | 4 | 0,072 |
| *PRKACB* | 5567 | Hypo | 4 | 0,054 |
| *TNFRSF1B* | 7133 | Hyper | 4 | 0,078 |
| *CD59* | 966 | Hyper | 3 | 0,085 |
| *CDC26* | 246184 | Hyper | 3 | 0,054 |
| *CDH11* | 1009 | Hyper | 3 | 0,054 |
| *CDH12* | 1010 | Mixed | 3 | 0,074 |
| *CDH13* | 1012 | Mixed | 3 | 0,084 |
| *CDH24* | 64403 | Hyper | 3 | 0,051 |
| *FANCI* | 55215 | Hypo | 3 | 0,051 |
| *KCNQ1* | 3784 | Hyper | 3 | 0,056 |
| *PAK2* | 5062 | Hyper | 3 | 0,059 |
| *RUNX3* | 864 | Hyper | 3 | 0,168 |
| *ATP8B4* | 79895 | Mixed | 2 | 0,130 |
| *CEACAM7* | 1087 | Hyper | 2 | 0,072 |
| *COL18A1* | 80781 | Hyper | 2 | 0,086 |
| *COL4A2* | 1284 | Hypo | 2 | 0,090 |
| *FAAP20* | 199990 | Hyper | 2 | 0,056 |
| *FANCA* | 2175 | Hypo | 2 | 0,056 |
| *HLA-B* | 3106 | Hyper | 2 | 0,077 |
| *HLA-C* | 3107 | Hyper | 2 | 0,087 |
| *HLA-G* | 3135 | Hyper | 2 | 0,054 |
| *IGFBP7* | 3490 | Hypo | 2 | 0,050 |
| *ITGB7* | 3695 | Hypo | 2 | 0,063 |
| *KIF18B* | 146909 | Hypo | 2 | 0,052 |
| *LIMD1* | 8994 | Hyper | 2 | 0,086 |
| *NOS3* | 4846 | Hypo | 2 | 0,059 |
| *NTNG1* | 22854 | Hypo | 2 | 0,059 |
| *OPCML* | 4978 | Mixed | 2 | 0,134 |
| *PPP2R5C* | 5527 | Hypo | 2 | 0,054 |
| *TNFSF14* | 8740 | Hyper | 2 | 0,122 |
| *ADRA1A* | 148 | Hyper | 1 | 0,050 |
| *ALDH1A1* | 216 | Hypo | 1 | 0,052 |
| *ATG101* | 60673 | Hyper | 1 | 0,101 |
| *CAMK1G* | 57172 | Hyper | 1 | 0,120 |
| *CCL16* | 6360 | Hyper | 1 | 0,094 |
| *CDKAL1* | 54901 | Hypo | 1 | 0,059 |
| *EXOC2* | 55770 | Hyper | 1 | 0,050 |
| *GAD1* | 2571 | Hyper | 1 | 0,068 |
| *GRIN2B* | 2904 | Hypo | 1 | 0,055 |
| *HDAC7* | 51564 | Hypo | 1 | 0,050 |
| *INPP5D* | 3635 | Hypo | 1 | 0,055 |
| *KCNAB1* | 7881 | Hyper | 1 | 0,052 |
| *KIF26B* | 55083 | Hyper | 1 | 0,058 |
| *LATS1* | 9113 | Hypo | 1 | 0,067 |
| *MEIS1* | 4211 | Hypo | 1 | 0,057 |
| *MYO5A* | 4644 | Hyper | 1 | 0,082 |
| *OAZ3* | 51686 | Hyper | 1 | 0,055 |
| *P2RY12* | 64805 | Hyper | 1 | 0,061 |
| *PRKAG2* | 51422 | Hypo | 1 | 0,070 |
| *SPTBN1* | 6711 | Hypo | 1 | 0,063 |
| *TRIO* | 7204 | Hypo | 1 | 0,068 |
| *ADGRV1* | 84059 | Hyper | 0 | 0,053 |
| *APC2* | 10297 | Hyper | 0 | 0,061 |
| *BIVM-ERCC5* | 100533467 | Hyper | 0 | 0,059 |
| *C9orf47* | 286223 | Hyper | 0 | 0,087 |
| *CACNG1* | 786 | Hyper | 0 | 0,053 |
| *CBFA2T3* | 863 | Hypo | 0 | 0,080 |
| *CELSR1* | 9620 | Hyper | 0 | 0,103 |
| *CHST11* | 50515 | Hypo | 0 | 0,074 |
| *COX6A2* | 1339 | Hypo | 0 | 0,073 |
| *CPA1* | 1357 | Hyper | 0 | 0,062 |
| *CSGALNACT1* | 55790 | Hypo | 0 | 0,102 |
| *CXXC1* | 30827 | Hyper | 0 | 0,056 |
| *CYP51A1* | 1595 | Hypo | 0 | 0,052 |
| *DSC3* | 1825 | Hyper | 0 | 0,065 |
| *ENTPD3* | 956 | Hyper | 0 | 0,066 |
| *GALNT13* | 114805 | Hypo | 0 | 0,065 |
| *GALNTL6* | 442117 | Hyper | 0 | 0,139 |
| *GULP1* | 51454 | Hypo | 0 | 0,051 |
| *MAEA* | 10296 | Hypo | 0 | 0,081 |
| *OS9* | 10956 | Hyper | 0 | 0,059 |
| *PDHX* | 8050 | Hyper | 0 | 0,067 |
| *PHF21A* | 51317 | Hyper | 0 | 0,062 |
| *PIGQ* | 9091 | Hyper | 0 | 0,090 |
| *ROR2* | 4920 | Hyper | 0 | 0,093 |
| *RPL27A* | 6157 | Hyper | 0 | 0,064 |
| *SPG7* | 6687 | Hypo | 0 | 0,078 |
| *XYLT1* | 64131 | Hypo | 0 | 0,067 |

**Table S9.** Lists of shared DMGs from the network analyses, and their corresponding direction of methylation. DMGs were defined as genes with at least one DMC, and the direction of methylation depended on the direction of methylation for all DMCs in the gene. Hypomethylation is defined as > -5% mean methylation difference (MMD), hypermethylation as > +5% MMD and mixed methylation genes contained DMPs of both directions. DMG – differentially methylated gene, DMC – differentially methylated CpG, L – *Lactobacillus reuteri*, MMD – mean methylation difference, ω – ω-3 fatty acids, P – placebo.

|  | Group comparison | | |
| --- | --- | --- | --- |
| Gene symbol | **Lω-PP** | **LP-PP** | **Pω-PP** |
| *CDH13* | Hyper | Hyper | Mixed |
| *CEACAM7* | Hyper | Hyper | Hyper |
| *CSGALNACT1* | Hypo | Mixed | Hypo |
| *HLA-B* | Hyper | Hyper | Hyper |
| *ITGB7* | Hypo | Hyper | Hypo |
| *KIF26B* | Hyper | Hypo | Hyper |
| *LAMC1* | Hyper | Hyper | Mixed |
| *NTNG1* | Hypo | Hypo | Hypo |
| *PHF21A* | Hyper | Hyper | Hyper |
| *RUNX3* | Hyper | Hyper | Hyper |
|  | **Group comparison** | | |
| Gene symbol | **Lω-PP** | **LP-PP** |  |
| *ADCY9* | Mixed | Hyper |  |
| *ADRB3* | Hyper | Hyper |  |
| *ASAP1* | Hyper | Hyper |  |
| *CACNA1A* | Hyper | Hyper |  |
| *CLOCK* | Hypo | Hypo |  |
| *CYB5R3* | Hyper | Hyper |  |
| *GALNT7* | Hyper | Hyper |  |
| *GALNT9* | Hypo | Hyper |  |
| *GP6* | Hyper | Hyper |  |
| *GPR31* | Hyper | Hyper |  |
| *HTR2B* | Hyper | Hyper |  |
| *INSIG1* | Hyper | Hyper |  |
| *MKLN1* | Hypo | Hyper |  |
| *PDIA6* | Hyper | Hyper |  |
| *PROK2* | Hyper | Hypo |  |
| *PRTN3* | Hypo | Hypo |  |
| *RCHY1* | Hyper | Hyper |  |
| *RHOH* | Hyper | Hyper |  |
| *RTN4* | Hyper | Hypo |  |
| *SLC2A3* | Hypo | Hypo |  |
| *STK10* | Hypo | Hypo |  |
| *TMPRSS6* | Hypo | Hypo |  |
| *TRPC6* | Hyper | Hyper |  |
| *WFS1* | Hyper | Hyper |  |
|  | **Group comparison** | | |
| Gene symbol | **Lω-PP** | **Pω-PP** |  |
| *ALDH1A1* | Hypo | Hypo |  |
| *ATG101* | Hyper | Hyper |  |
| *C9orf47* | Hyper | Hyper |  |
| *CCL16* | Hyper | Hyper |  |
| *CDH12* | Hypo | Mixed |  |
| *CDKN1A* | Hyper | Hyper |  |
| *GALNT13* | Mixed | Hypo |  |
| *GRB2* | Hyper | Hypo |  |
| *MEIS1* | Hypo | Hypo |  |
| *MYO5A* | Hyper | Hyper |  |
| *OPCML* | Hypo | Mixed |  |
| *OS9* | Hyper | Hyper |  |
| *PAK2* | Hyper | Hyper |  |
| *PRKACB* | Hypo | Hypo |  |
|  | **Group comparison** | | |
| Gene symbol | **LP-PP** | **Pω-PP** |  |
| *ADRA1A* | Hyper | Hyper |  |
| *CBFA2T3* | Hypo | Hypo |  |
| *CCNB1* | Hyper | Hyper |  |
| *DSC3* | Hyper | Hyper |  |
| *ENTPD3* | Hyper | Hyper |  |
| *GALNTL6* | Hypo | Hyper |  |
| *ITGB4* | Hyper | Hyper |  |
| *KCNQ1* | Hyper | Hyper |  |
| *ROR2* | Hyper | Hyper |  |
| *XYLT1* | Hypo | Hypo |  |

**Table S10.** A table of genes overlapping between comparisons of DMCs in differential DNA methylation analyses (Lω, LP and Pω) to the publicly available allergy data set.

| **Genes overlapping between** | | | | | | | |
| --- | --- | --- | --- | --- | --- | --- | --- |
| **All comparisons (Lω, LP and Pω)** | | **Lω *vs.* LP** | | **Lω *vs.* Pω** | | **LP *vs.* Pω** | |
| cg14060519 | *MYOF* | cg14060519 | *MYOF* | cg12133451 | *NA* | cg14060519 | *MYOF* |
| cg05733780 | *C6orf123* | cg15205559 | *NA* | cg14060519 | *MYOF* | cg27474175 | *ZFAND6* |
| cg10473311 | *PTPRN2* | cg16786808 | *EPHA4* | cg05830791 | *NA* | cg05733780 | *C6orf123* |
|  |  | cg05733780 | *C6orf123* | cg26394940 | *C22orf26* | cg10473311 | *PTPRN2* |
|  |  | cg10473311 | *PTPRN2* | cg05733780 | *C6orf123* | cg11953088 | *NA* |
|  |  | cg00330059 | *ODZ3* | cg10473311 | *PTPRN2* | cg17740434 | *MIR548N* |
|  |  | cg02390801 | *CCDC33* | cg05482498 | *NA* | cg26466508 | *FILIP1* |
|  |  | cg03035115 | *NA* | cg07327178 | *NA* |  |  |
|  |  | cg06804823 | *NA* | cg09230756 | *NA* |  |  |
|  |  | cg11479811 | *PP14571* | cg14743812 | *NA* |  |  |
|  |  | cg27306802 | *MEGF6* | cg14950193 | *C7orf63* |  |  |
|  |  |  |  | cg19421526 | *CRTAC1* |  |  |
|  |  |  |  | cg19735538 | *C6orf227* |  |  |
|  |  |  |  | cg27307810 | *LOC154822* |  |  |

CpG position identifiers and gene names are reported (when applicable). NA – not applicable.

**Table S11.** Cell type proportions in the included CD4+ T cell samples, as determined using the Houseman method.

| **Intervention group** | **Lω** | **LP** | **Pω** | **PP** |
| --- | --- | --- | --- | --- |
| CD8+ T cells | 1 (0-3) | 2 (1-4) | 2 (1-3) | 2 (1-3) |
| CD4+ T cells | 96 (94-98) | 94 (93-96) | 95 (92-98) | 96 (94-97) |
| NK cells | 0 (0-0) | 0 (0-0) | 0 (0-0) | 0 (0-0) |
| B cells | 0 (0-0) | 1 (0-1) | 0 (0-0) | 0 (0-0) |
| Monocytes | 0 (0-0) | 0 (0-0) | 0 (0-0) | 0 (0-0) |
| Granulocytes | 1 (1-3) | 3 (2-3) | 2 (1-4) | 2 (1-3) |

The proportions of each cell type are expressed as medians with interquartile
ranges in brackets. L – *Lactobacillus reuteri*, ω – ω-3 fatty acids, P – placebo.
